# Artificial intelligence for precision oncology: AI-HOPE-MAPK uncovers clinically actionable MAPK alterations in colorectal cancer

**DOI:** 10.1101/2025.06.11.25329446

**Authors:** Ei-Wen Yang, Brigette Waldrup, Enrique Velazquez-Villarreal

**Author notes:** Corresponding Author –.

## Abstract

**Background:** The emergence of early-onset colorectal cancer (EOCRC), particularly among populations with disproportionate health burdens, has exposed critical gaps in our understanding of age– and ancestry-specific oncogenic processes. Aberrations in the mitogen-activated protein kinase (MAPK) pathway—encompassing KRAS, NRAS, BRAF, MAP2K1, MAPK3, and others—are central to tumorigenesis, yet their distribution, prognostic impact, and therapeutic implications in EOCRC remain poorly defined.

**Methods:** We developed AI-HOPE-MAPK, a conversational artificial intelligence platform built upon a fine-tuned biomedical large language model (LLaMA 3) to enable real-time, natural language-driven analysis of clinical and genomic data. The platform integrates harmonized datasets from TCGA, cBioPortal, and AACR Project GENIE and supports cohort construction, mutation frequency comparisons, survival modeling, and odds ratio estimation across diverse subgroups. We applied AI-HOPE-MAPK to characterize MAPK pathway alterations in colorectal cancer by age, race/ethnicity, microsatellite instability (MSI), tumor mutational burden (TMB), treatment history, and anatomical site.

**Results:** AI-HOPE-MAPK identified significant age– and ancestry-specific patterns in MAPK alterations. NF1 and MAP2K1 mutations were enriched in H/L EOCRC cases, with NF1 mutations reaching statistical significance (p = 0.045). Comparative analyses revealed elevated MAPK3 and NF1 mutation frequencies in Hispanic/Latino and non-Hispanic White EOCRC patients. MAP2K1-mutated stage I–III tumors exhibited worse survival and lower FOLFOX treatment rates (p = 0.0447 and p = 0.015, respectively). Kaplan-Meier and odds ratio analyses showed that younger patients with low-TMB MAPK-altered tumors had significantly poorer outcomes and were overrepresented in aggressive molecular subtypes. Furthermore, MAPK alterations conferred significantly reduced survival in microsatellite-stable (MSS) tumors but not in MSI-H cases, underscoring context-specific prognostic effects. Anatomical stratification showed site-specific impacts of AKT1 mutations, with poorer outcomes in colon versus rectal cancers.

**Conclusions:** AI-HOPE-MAPK offers a transformative framework for AI-driven precision oncology by enabling intuitive, scalable, and population-aware exploration of MAPK pathway dysregulation in colorectal cancer. The platform revealed clinically actionable differences in mutation prevalence, survival outcomes, and treatment access, particularly among patients with disproportionate health burdens. By combining language-based user interfaces with rigorous genomic analytics, AI-HOPE-MAPK accelerates biomarker discovery, supports equitable research practices, and advances the implementation of precision oncology across multiple patient populations.

## Introduction

Colorectal cancer (CRC) remains a major global health challenge, ranking as the third most common malignancy and the second leading cause of cancer-related deaths worldwide [1,2]. Although CRC incidence in older adults has plateaued or declined in high-income countries, the global rise in early-onset colorectal cancer (EOCRC)— defined as diagnosis before age 50—has garnered increasing attention [3–6]. In the United States, this trend is especially pronounced among Hispanic/Latino (H/L) populations, who face the most significant increase in EOCRC incidence as well as disproportionately high mortality rates compared to non-Hispanic White (NHW) individuals [7–12]. These disparities are compounded by later-stage diagnoses resulting from current screening guidelines that historically initiated routine surveillance at age 50 [13].

Beyond delayed detection, EOCRC displays distinct molecular and immunological features when compared to late-onset CRC (LOCRC). Studies have identified a higher prevalence of microsatellite instability (MSI), increased tumor mutation burden, and overexpression of immune checkpoint proteins such as PD-L1 in EOCRC cases [12–17]. Epigenetic modifications, such as LINE-1 hypomethylation, have also emerged as potential molecular signatures distinguishing EOCRC from LOCRC [18–21]. These findings suggest that EOCRC may be driven by unique, age– and ancestry-specific oncogenic mechanisms that remain poorly understood in racially and ethnically diverse populations.

Among the most critical pathways implicated in CRC development is the mitogen-activated protein kinase (MAPK) signaling cascade. Comprising ERK, BMK-1, JNK, and p38 branches, the MAPK pathway orchestrates key cellular processes such as proliferation, differentiation, and apoptosis [22–25]. In CRC, aberrant MAPK activation— often through RAS and BRAF mutations—leads to uncontrolled tumor growth, metastasis, and resistance to therapy [26–28]. Importantly, MAPK reactivation via EGFR signaling has been shown to confer resistance to BRAF inhibitors like vemurafenib, underscoring the clinical relevance of pathway-specific therapeutic vulnerabilities [20].

Despite extensive research into MAPK-driven CRC biology [29–33], its role in shaping EOCRC disparities—particularly among H/L individuals—remains largely unexplored.

To address this critical knowledge gap, we developed AI-HOPE-MAPK, a conversational artificial intelligence platform designed to identify, analyze, and interpret MAPK pathway alterations in CRC using clinical and genomic data. AI-HOPE-MAPK integrates publicly available datasets, supports natural language-driven analysis, and enables population-aware interrogation of pathway-specific alterations. Here, we apply AI-HOPE-MAPK to characterize the molecular landscape of MAPK signaling in EOCRC across diverse patient groups. By linking genomic alterations to clinical outcomes, this study aims to uncover actionable targets and support the development of tailored precision oncology strategies for different communities.

## Methods

AI-HOPE-MAPK was developed as an artificial intelligence–powered system designed to translate natural language inquiries into rigorous, clinically meaningful genomic analyses, with a specific focus on alterations in the MAPK signaling pathway in CRC. The system is built upon a fine-tuned biomedical large language model (LLM) based on the LLaMA 3 architecture, which enables semantic understanding of user queries and automatic generation of analytical workflows. Researchers can input complex, multi-layered questions—for example, assessing survival outcomes associated with specific mutations in microsatellite-stable, early-onset, ethnicity-specific CRC cases—and receive comprehensive statistical outputs without the need for computational programming expertise.

The platform integrates harmonized clinical and genomic data from cBioPortal and the AACR Project GENIE. These datasets were filtered to include colorectal adenocarcinomas with complete annotations for somatic mutations, copy number alterations, microsatellite instability (MSI) status, tumor site, clinical stage, patient race/ethnicity, age at diagnosis, treatment history, and survival time. A standardized preprocessing pipeline was employed to normalize patient identifiers, resolve inconsistent annotations, and map all gene alterations to predefined MAPK pathway components, including KRAS, NRAS, BRAF, MAP2K1, MAPK1, and MAPK14. Tumors were stratified by anatomical location (left colon, right colon, rectum), MSI phenotype, and racial or ethnic background, with particular emphasis on distinguishing EOCRC (diagnosed before age 50) from later-onset disease.

Upon receiving a user query, AI-HOPE-MAPK applies a semantic parsing engine to extract analytical intent, identify the genes and pathways involved, and apply relevant cohort filters. This automated translation process initiates a statistical pipeline tailored to the nature of the question. The system supports frequency analysis of mutations and copy number alterations across clinical subgroups using chi-square or Fisher’s exact tests, calculates odds ratios and 95% confidence intervals to assess enrichment of alterations by demographic or molecular category, and conducts time-to-event analyses including Kaplan-Meier estimation, log-rank testing, and Cox proportional hazards regression. Pathway interaction modeling is also enabled, including co-occurrence and mutual exclusivity analysis with other relevant oncogenic pathways such as TP53, TGF-β, and PI3K.

To validate the system’s performance, we reproduced known associations between MAPK pathway mutations and clinical outcomes in CRC, including the poor prognosis of BRAF V600E mutations and the differential impact of RAS mutations in microsatellite-stable tumors. The platform was benchmarked against publicly available bioinformatics tools such as cBioPortal and UCSC Xena, with evaluation criteria including cohort construction accuracy, response time, and clarity of output. In all test cases, AI-HOPE-MAPK demonstrated strong reproducibility and superior flexibility in handling queries that involved multiple intersecting variables, including ethnicity, tumor location, and MSI status.

All analytical outputs are returned as high-resolution visualizations and narrative summaries. These include Kaplan-Meier survival curves annotated with p-values and sample sizes, forest plots of estimated effect sizes, co-mutation heatmaps, and subgroup-specific mutation frequency charts. Result tables are exportable in multiple formats for downstream use in manuscripts, presentations, or supplemental materials. The conversational interface also supports iterative querying, enabling users to refine hypotheses or generate stratified comparisons across subgroups in real time.

By bridging the gap between biomedical language understanding and pathway-specific clinical-genomic analysis, AI-HOPE-MAPK provides a powerful, accessible tool for investigating the molecular underpinnings of CRC and identifying disparities in disease presentation and outcomes across diverse populations.

## Results

AI-HOPE-MAPK provides an intuitive, conversation-based framework for investigating clinically relevant alterations in the MAPK signaling pathway among CRC patients from multiple populations. Leveraging natural language inputs, the system constructs cohort-specific analyses spanning mutation frequency comparisons, survival modeling, and odds ratio testing, while incorporating key clinical variables such as age, ethnicity, tumor site, and treatment history. The platform enables hypothesis-driven exploration of CRC disparities, particularly among early-onset and H/L subgroups, through real-time, data-integrated insights.

### Age-associated molecular trends in population-specific CRC

AI-HOPE-MAPK was first applied to explore molecular distinctions between EOCRC and LOCRC in H/L patients. Although infrequent, ACVR1 mutations appeared more often in EOCRC H/L cases (1.96%) than in LOCRC H/L counterparts (0.49%), yielding an odds ratio of 4.06 (95% CI: 0.418–39.416; p = 0.425) (Figure 2). While not statistically significant, this pattern hints at age-specific enrichment requiring validation. In contrast, NF1 mutations showed a statistically significant difference, with a 10.46% prevalence in EOCRC H/L samples compared to 4.41% in LOCRC H/L (OR = 2.53; 95% CI: 1.087–5.893; p = 0.045) (Figure S1), suggesting a potentially pathogenic role in early-onset disease.

**Figure 1.**
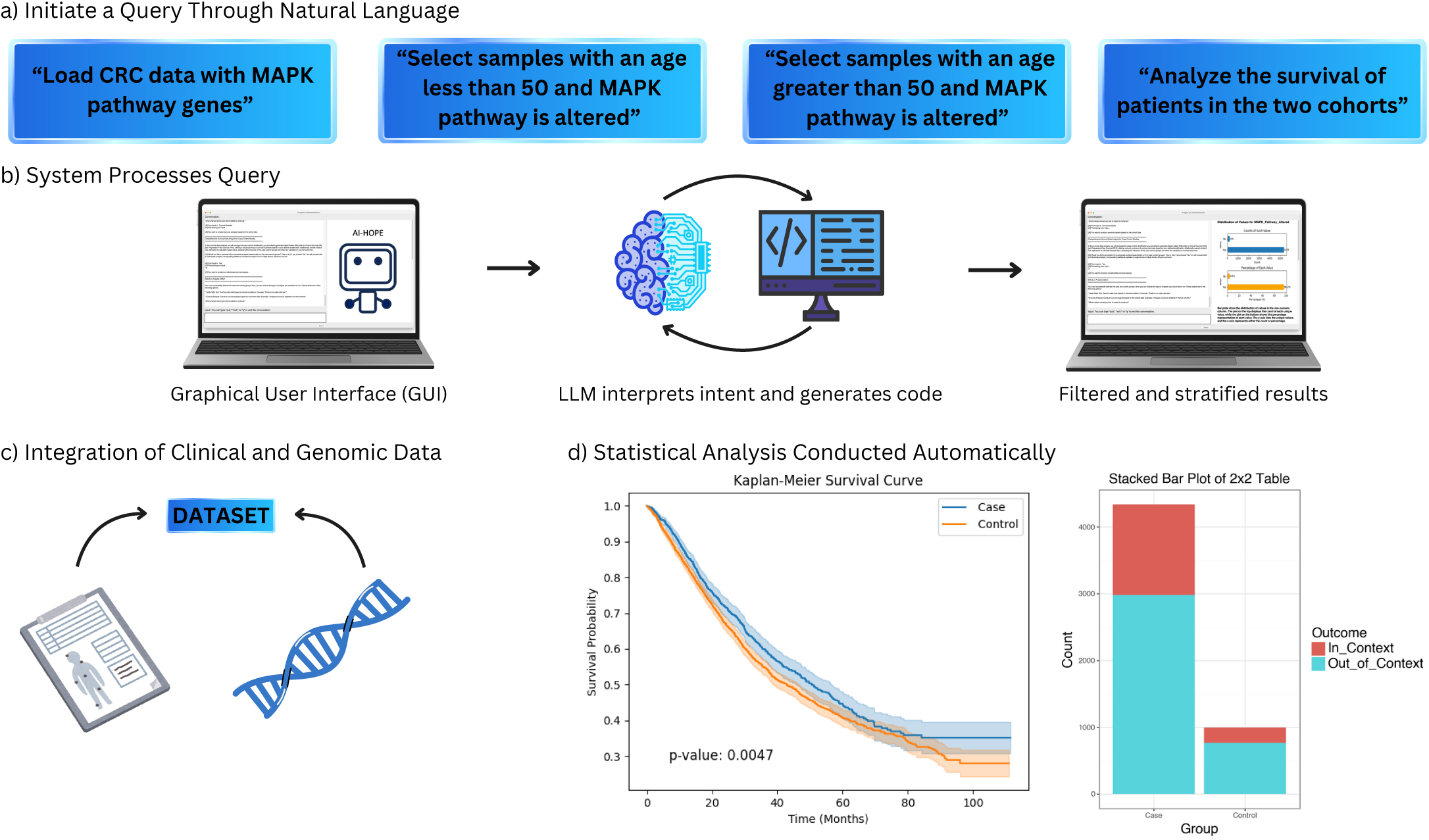
AI-HOPE-MAPK workflow for discovering clinically relevant MAPK alterations in colorectal cancer. This figure presents a visual summary of the AI-HOPE-MAPK system, an interactive artificial intelligence platform designed to explore MAPK pathway alterations in colorectal cancer (CRC) through natural language queries. (a) Users begin by inputting intuitive prompts such as loading CRC datasets and specifying age– or mutation-based criteria—e.g., selecting patients under or over 50 years old with MAPK pathway alterations. (b) The system processes these queries through a graphical user interface (GUI) powered by a large language model (LLM), which translates user intent into executable code. (c) Clinical and genomic data are integrated in real time, enabling immediate generation of stratified case-control cohorts. (d) Automated statistical analyses—including survival comparisons—are conducted and visualized, delivering actionable insights into age-stratified MAPK pathway alterations relevant to CRC prognosis and therapeutic strategies.

**Figure 2.**
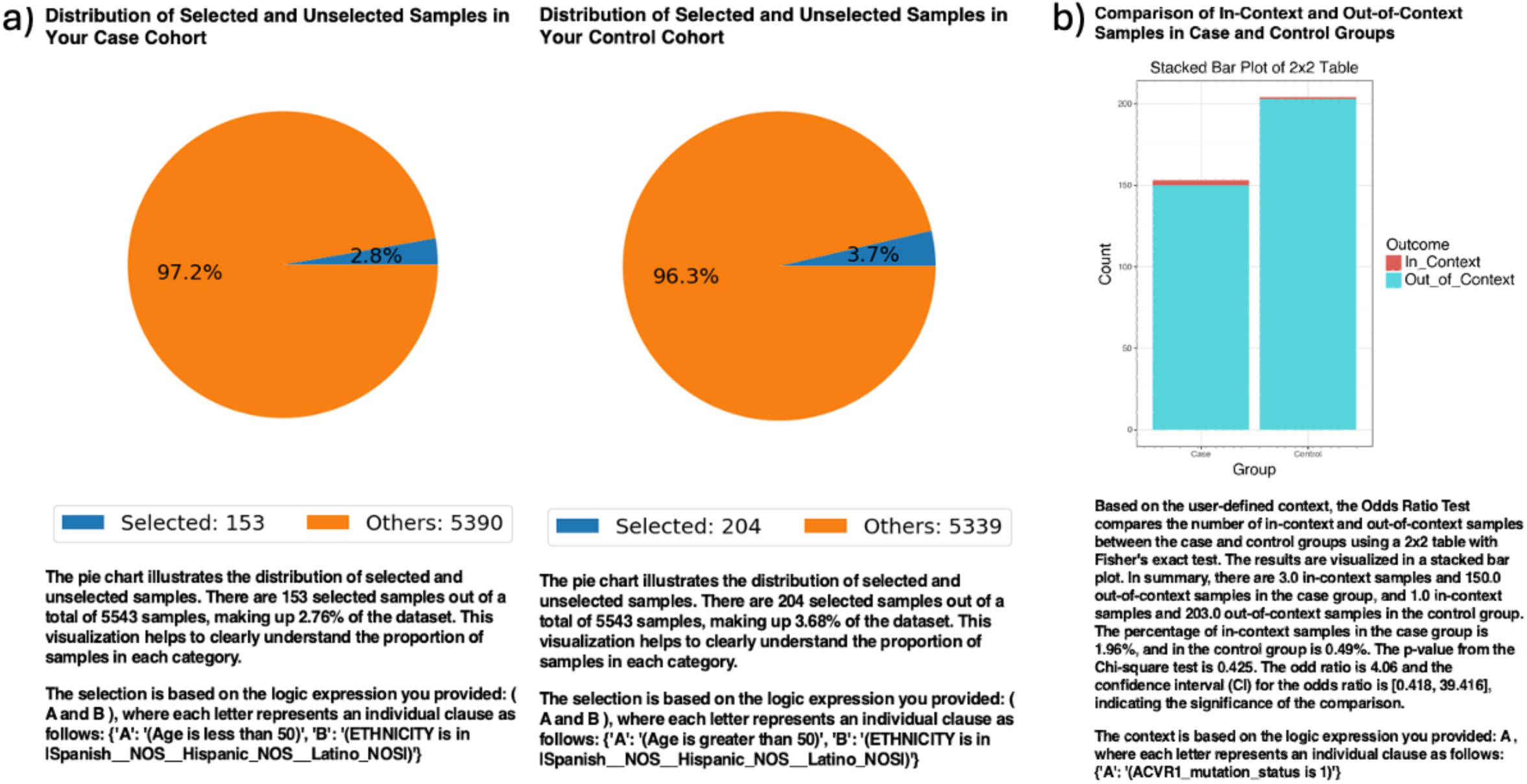
AI-HOPE-MAPK analysis of ACVR1 mutation frequency in early-onset versus late-onset Hispanic/Latino colorectal cancer patients. a) Pie charts illustrate the proportion of selected samples in each cohort following natural language–based filtering by AI-HOPE-MAPK. The case cohort consists of 153 early-onset colorectal cancer (EOCRC) samples from Hispanic/Latino patients under age 50, comprising 2.76% of the dataset. The control cohort includes 204 late-onset colorectal cancer (LOCRC) samples from Hispanic/Latino patients over age 50, representing 3.68% of the dataset. These visualizations depict the relative sample representation used for downstream comparative analyses. b) A 2×2 odds ratio analysis evaluates the distribution of ACVR1 mutations between the EOCRC HL and LOCRC HL groups. The stacked bar plot shows the number of samples with (“In_Context”) and without (“Out_of_Context”) ACVR1 mutations in each group. ACVR1 mutations were observed in 1.96% of EOCRC HL samples and 0.49% of LOCRC HL samples. The calculated odds ratio was 4.06 (95% CI: [0.418, 39.416]) with a p-value of 0.425, indicating no statistically significant difference. While not conclusive, this trend suggests a potential enrichment of ACVR1 mutations in early-onset Hispanic/Latino CRC, warranting further validation in larger datasets.

MAP2K1 alterations—although rare—were restricted almost entirely to the EOCRC H/L group (2.61% vs. 0.24% in LOCRC H/L), producing an infinite odds ratio (95% CI: 0.575–208.75; p = 0.07) due to absence in the older cohort (Figure S2). In contrast, BRAF mutations were more prominent in LOCRC H/L samples (17.65%) than EOCRC H/L (5.23%) and achieved statistical significance (OR = 3.88; 95% CI: 1.749–8.624; p = 0.001) (Figure S3), underscoring the potential age-dependent divergence in MAPK pathway biology.

### Ancestry-informed differences in MAPK gene alterations

To assess population-level disparities, AI-HOPE-MAPK compared mutation patterns between H/L and NHW EOCRC patients. While AKT1 mutation rates showed a modest, non-significant increase in H/L cases (3.27% vs. 1.52%, OR = 2.19; 95% CI: 0.795– 6.013; p = 0.222) (Figure S4), MAPK3 mutations were significantly more common in the H/L group (2.61% vs. 0.63%; OR = 4.26; 95% CI: 1.232–14.715; p = 0.043) (Figure S5).

Similarly, NF1 mutations occurred at a higher rate in H/L patients (10.46%) compared to NHW (5.37%) with statistical significance (OR = 2.06; 95% CI: 1.153–3.673; p = 0.021) (Figure S6). PDGFRB mutations showed a similar trend (3.92% vs. 1.97%, OR = 2.03; 95% CI: 0.81–5.092; p = 0.212), though did not reach significance (Figure S7). These observations reinforce the relevance of ancestry-stratified analyses in identifying population-specific MAPK alterations.

### Therapeutic disparities and treatment context

AI-HOPE-MAPK further examined age-specific responses to standard FOLFOX chemotherapy in H/L CRC patients. Kaplan-Meier analysis demonstrated significantly worse survival among early-onset cases compared to late-onset patients (p = 0.0047) (Figure 3). Odds ratio testing revealed that H/L individuals comprised a disproportionately higher fraction of the early-onset group (10.29%) than the late-onset group (5.35%) (OR = 1.957; 95% CI: 1.534–2.497; p < 0.001), suggesting a potential convergence of ancestry and age-related vulnerability in CRC.

**Figure 3.**
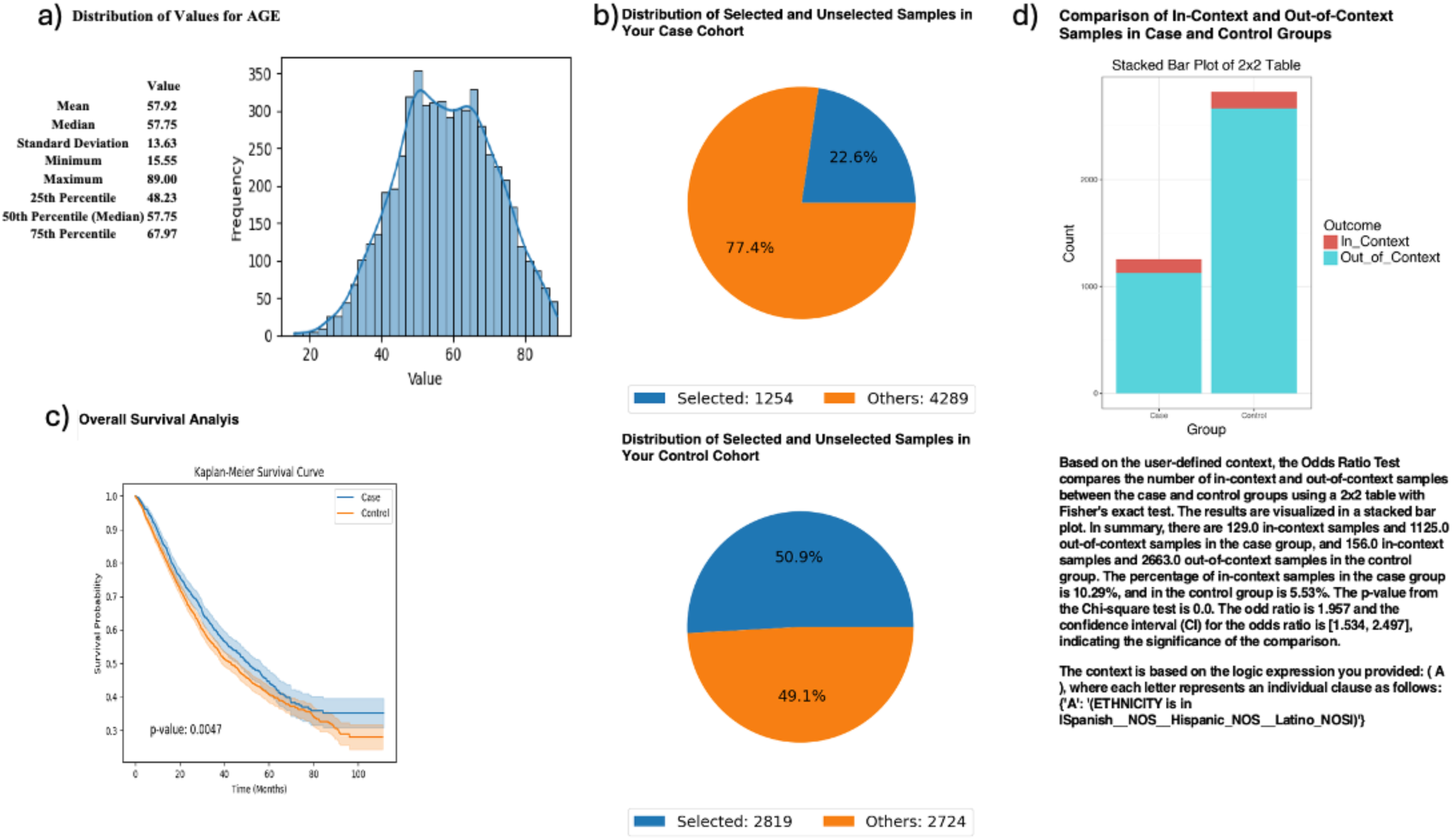
AI-HOPE-MAPK analysis of age-stratified Hispanic/Latino colorectal cancer patients treated with FOLFOX chemotherapy. This figure demonstrates the use of AI-HOPE-MAPK to investigate age-related survival patterns and ethnic disparities among colorectal cancer (CRC) patients treated with the FOLFOX regimen (Fluorouracil, Leucovorin, Oxaliplatin). a) A histogram illustrates the distribution of patient age across the full dataset, showing a mean of 57.92 years and a median of 57.75, supporting the stratification of case (age <50) and control (age >50) cohorts. b) Pie charts display the proportion of selected and unselected samples following natural language query filtering. The early-onset (case) cohort includes 1,254 patients (22.6% of the dataset), while the late-onset (control) cohort includes 2,819 patients (50.9%). c) Kaplan-Meier survival analysis compares overall survival between the two age groups. The early-onset cohort demonstrated significantly worse survival compared to the late-onset group, with a p-value of 0.0047, suggesting potential age-related differences in treatment response or disease biology in patients receiving FOLFOX. d) A 2×2 odds ratio analysis evaluates the distribution of Hispanic/Latino ethnicity across the cohorts. Hispanic/Latino patients represented 10.29% of the early-onset group and 5.35% of the late-onset group, yielding an odds ratio of 1.957 (95% CI: 1.534–2.497) and a statistically significant p-value of 0.0. These results highlight the disproportionate representation of Hispanic/Latino patients in the early-onset cohort and support further investigation into ancestry-specific treatment outcomes and MAPK-related therapeutic vulnerabilities in FOLFOX-treated CRC.

### Survival differences by MSI and MAPK interaction

AI-HOPE-MAPK was used to assess how MAPK alterations intersect with microsatellite instability (MSI) status in FOLFOX-treated patients. Although age-based enrichment of younger patients did not differ significantly between MSS and MSI-H groups (OR = 1.206; 95% CI: 0.893–1.629; p = 0.251), survival analysis showed a striking disadvantage for microsatellite-stable (MSS) tumors with MAPK alterations compared to their MSI-H counterparts (p < 0.0001) (Figure 4). This divergence underscores the importance of MSI context in stratifying risk among MAPK-driven CRC.

**Figure 4.**
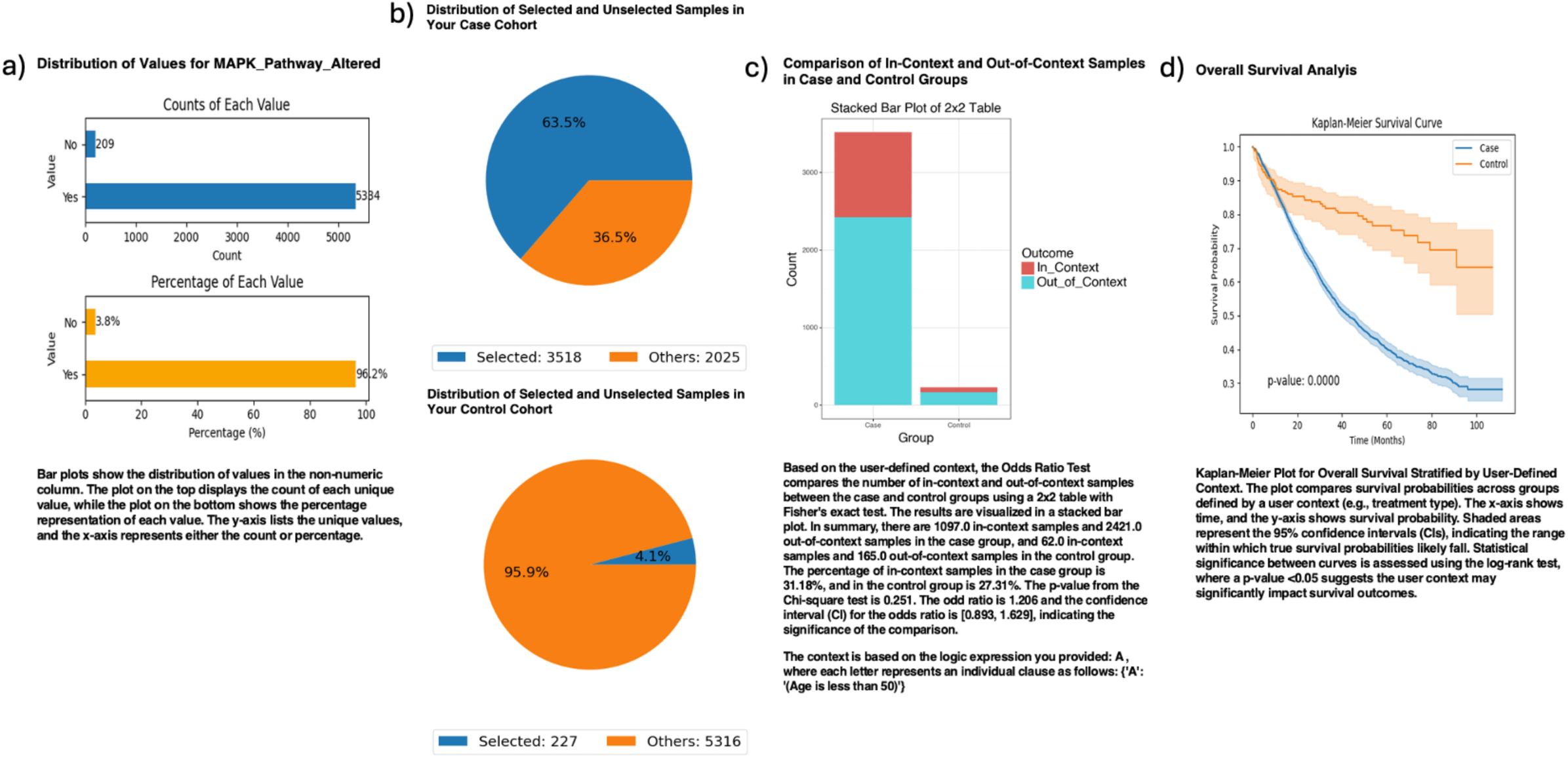
AI-HOPE-MAPK analysis of survival and enrichment patterns in MAPK-altered colorectal cancer stratified by MSI status and chemotherapy exposure. This figure showcases the application of AI-HOPE-MAPK to investigate survival outcomes and age-based enrichment in microsatellite-stable (MSS) versus microsatellite-instability-high (MSI-H) colorectal cancer (CRC) patients with MAPK pathway alterations who were treated with FOLFOX chemotherapy (Fluorouracil, Leucovorin, Oxaliplatin). a) Bar plots illustrate the distribution and percentage of samples with MAPK pathway alterations across the dataset. Among all patients analyzed, 96.2% had MAPK pathway alterations, confirming a sufficient sample size for subgroup comparison. b) Pie charts summarize the selection of case and control cohorts based on MSI status. The case cohort, defined as MSS with MAPK alterations and FOLFOX treatment, includes 3,518 patients (63.5%), while the MSI-H control cohort includes 227 patients (4.1%). c) A 2×2 odds ratio analysis evaluates the age-based enrichment (under age 50) across the two groups. The proportion of younger patients in the MSS cohort was 31.8%, compared to 27.31% in the MSI-H cohort, resulting in an odds ratio of 1.206 (95% CI: 0.893–1.629; p = 0.251), indicating no statistically significant enrichment. d) Kaplan-Meier survival analysis reveals a striking difference in overall survival between MSS and MSI-H groups with MAPK alterations, showing significantly poorer survival in the MSS cohort (blue) compared to the MSI-H cohort (orange), with a p-value < 0.0001. The separation of the survival curves and the non-overlapping confidence intervals suggest that MSI status may strongly influence survival outcomes in MAPK-altered, FOLFOX-treated CRC patients. These findings underscore the need for MSI-aware treatment stratification and the potential prognostic value of MSI in MAPK-driven CRC.

### Anatomical and mutational site interactions

In a site-specific analysis, patients with AKT1-mutated colon adenocarcinomas showed significantly poorer overall survival compared to those with rectal tumors (p = 0.0483) (Figure 5), suggesting the modifying influence of tumor location on mutational prognosis.

**Figure 5.**
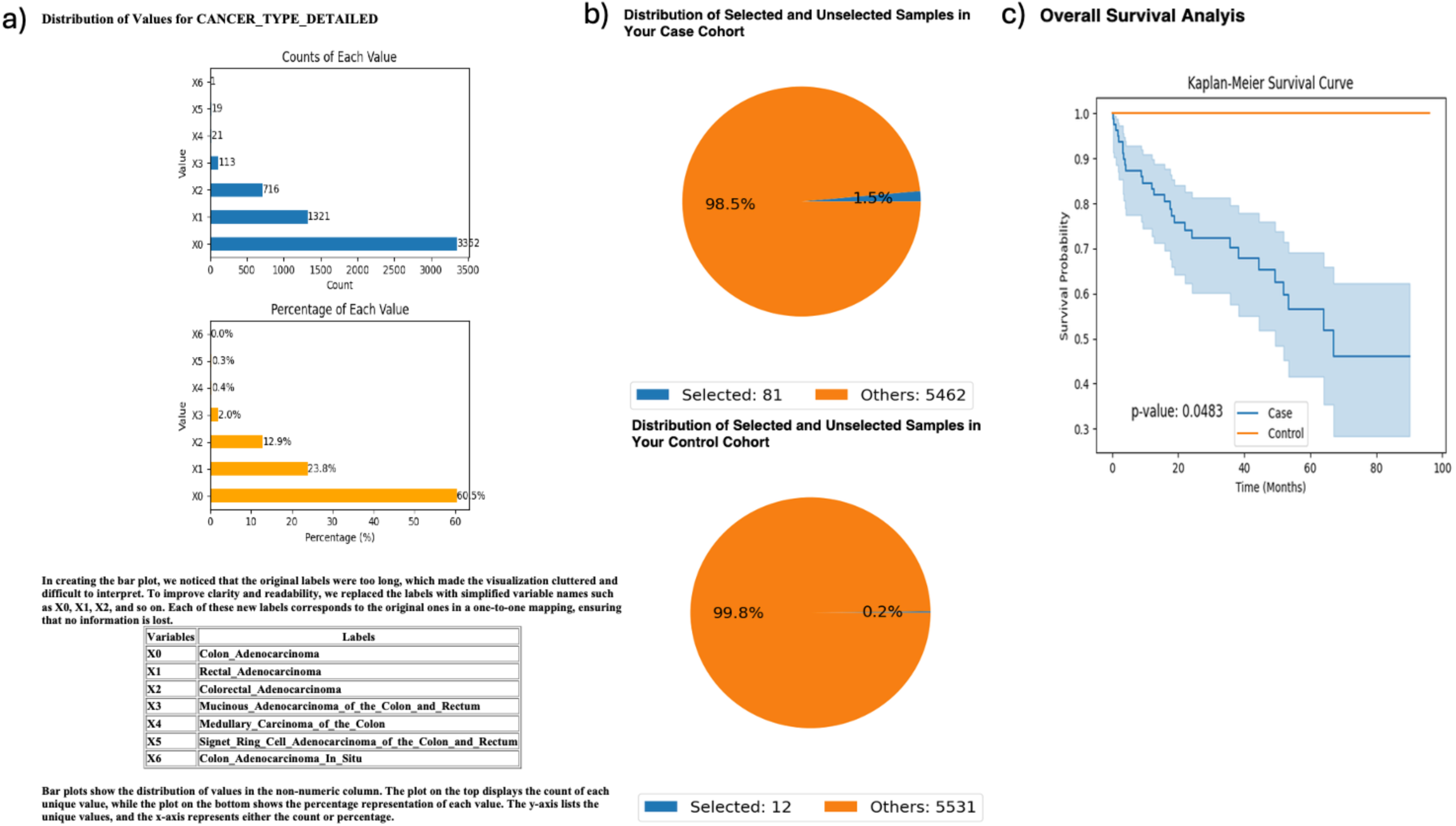
AI-HOPE-MAPK comparison of survival outcomes in AKT1-mutated colon versus rectal adenocarcinomas. This figure illustrates the use of AI-HOPE-MAPK to evaluate differences in clinical behavior among colorectal cancer (CRC) patients with AKT1 mutations, comparing those with colon adenocarcinoma (case group) to those with rectal adenocarcinoma (control group). a) Bar plots depict the overall distribution of cancer subtypes in the dataset using simplified category labels for readability. Colon adenocarcinoma (X0) accounted for the majority of cases (60.5%), followed by rectal adenocarcinoma (X1, 23.8%). b) Pie charts summarize the selection of AKT1-mutated cases by tumor location. Among 5,543 samples, 81 colon adenocarcinoma samples with AKT1 mutations (1.5%) were selected as the case group, while only 12 rectal adenocarcinoma samples with AKT1 mutations (0.2%) were selected as controls. c) Kaplan-Meier survival analysis compares overall survival between the two groups. Patients with AKT1-mutant colon adenocarcinoma exhibited significantly poorer survival than those with AKT1-mutant rectal adenocarcinoma, with a p-value of 0.0483. The survival curve indicates a clear divergence between the two tumor types, suggesting that tumor site may influence clinical outcomes among AKT1-mutant CRC patients. These findings highlight the utility of AI-HOPE-MAPK in dissecting mutation-specific, site-dependent survival patterns in colorectal cancer.

### TMB-defined molecular subtypes in MAPK-driven CRC

AI-HOPE-MAPK also examined tumor mutational burden (TMB) in relation to MAPK status. Patients with MAPK pathway alterations and low TMB (<10 mutations/Mb) showed significantly worse survival than high-TMB, MAPK-wildtype controls (p < 0.0001) (Figure S8). Odds ratio analysis revealed a significantly greater proportion of younger patients (<50) in the low-TMB MAPK-altered group (31.8%) than in controls (23.29%) (OR = 1.501; 95% CI: 1.278–1.762; p < 0.001), suggesting that EOCRC may harbor a unique, MAPK-driven, low-TMB molecular phenotype.

### Prognostic impact of MAP2K1 in early-stage disease

Finally, MAP2K1 mutations were evaluated in Stage I–III CRC patients. Mutant cases showed significantly poorer survival (p = 0.0447), and were less likely to receive FOLFOX (46.15% vs. 63.55%, OR = 0.491; 95% CI: 0.283–0.851; p = 0.015) (Figure S9). These findings suggest a prognostic and potentially treatment-modifying role for MAP2K1 in early-stage CRC.

## Discussion

In this study, we present the development and application of AI-HOPE-MAPK, a conversational artificial intelligence platform designed to uncover clinically relevant MAPK pathway alterations in CRC through integrative, population-aware analysis. Built upon a fine-tuned large language model, AI-HOPE-MAPK bridges natural language understanding with rigorous clinical-genomic computation, empowering researchers to stratify cohorts, identify mutation patterns, and evaluate survival outcomes without requiring programming expertise. By enabling real-time, multi-parameter interrogation of large-scale datasets, this platform addresses longstanding barriers in precision oncology—particularly those related to equity, accessibility, and representation in cancer genomics research.

Applying AI-HOPE-MAPK to EOCRC, with a focus H/L populations, yielded key insights into age– and ancestry-specific differences in MAPK signaling alterations. Notably, NF1 mutations were significantly more prevalent in H/L EOCRC cases compared to their late-onset counterparts, suggesting a potentially underappreciated role for NF1 in age-associated tumorigenesis. Although ACVR1 and MAP2K1 mutations did not reach statistical significance due to low frequency, their higher prevalence in EOCRC H/L patients indicates potential biological relevance that warrants further investigation. The consistent presence of these alterations in younger patients, in combination with the absence or lower frequency in late-onset cases, raises important questions about distinct oncogenic trajectories in EOCRC—particularly among underserved populations.

Our analysis also revealed significant ancestry-informed disparities in the mutation landscape. MAPK3 and NF1 mutations were significantly enriched in H/L EOCRC patients relative to NHW individuals, with MAPK3 displaying a four-fold increase in frequency. These results underscore the critical importance of ancestry-stratified analyses in genomic research, which have historically relied on predominantly NHW datasets. Without tools like AI-HOPE-MAPK, such population-specific mutation patterns—especially in genes less frequently studied outside canonical drivers like KRAS or BRAF—may remain undetected. These findings contribute to a growing body of evidence suggesting that certain MAPK pathway components may play divergent roles across racially and ethnically distinct CRC subtypes, and may offer novel therapeutic targets tailored to these groups.

In addition to uncovering mutation patterns, AI-HOPE-MAPK facilitated deeper exploration of therapeutic response and survival outcomes. We observed significantly worse survival in H/L EOCRC patients receiving FOLFOX chemotherapy compared to their late-onset counterparts, despite standardized treatment. This survival gap raises the possibility that younger patients, particularly from minoritized populations, may harbor distinct tumor biology—potentially characterized by immune evasion, chemotherapy resistance, or MAPK-mediated signaling resilience. These observations are consistent with earlier studies showing suboptimal responses in EOCRC despite aggressive disease management and highlight the need for more nuanced, biology-informed treatment strategies.

Further, our integrative analyses demonstrate the prognostic importance of combining MAPK pathway status with molecular markers such as microsatellite instability (MSI) and tumor mutational burden (TMB). Specifically, MAPK-altered tumors in microsatellite-stable (MSS) patients exhibited significantly worse survival than MSI-high (MSI-H) tumors, reinforcing MSI status as a critical contextual modifier of pathway-driven outcomes. Notably, CRC cases with low TMB and concurrent MAPK alterations were significantly enriched in younger patients and exhibited inferior prognosis, suggesting a unique molecular subtype of EOCRC that may be poorly responsive to both immunotherapy and conventional cytotoxic regimens. These findings highlight an urgent need for molecular profiling in young patients and support the development of alternative therapeutic approaches, such as MEK inhibitors or EGFR blockade tailored to specific MAPK pathway mutations.

Anatomical context also emerged as a relevant modifier of MAPK-associated prognosis. Patients with AKT1-mutated tumors in the colon experienced significantly poorer survival than those with similar mutations in rectal tumors, consistent with prior evidence suggesting that tumor site influences molecular evolution, treatment response, and disease progression. Such findings further underscore the need for integrated models of CRC biology that consider not only genomic alterations but also anatomical, demographic, and treatment-related variables.

The prognostic implications of MAP2K1 mutations in early-stage disease were particularly striking. Despite being uncommon, these mutations were associated with worse survival outcomes and a significantly lower likelihood of receiving FOLFOX chemotherapy. This observation suggests both biological aggressiveness and potential underutilization of standard treatment in this subgroup. Whether this reflects clinical decision-making based on comorbidities, physician perception of benefit, or other unmeasured social determinants remains unclear—but the ability of AI-HOPE-MAPK to rapidly surface such disparities offers a powerful framework for future clinical investigation.

Collectively, these findings demonstrate that AI-HOPE-MAPK not only recapitulates known CRC biology—such as BRAF enrichment in late-onset CRC—but also expands the landscape of actionable discoveries by uncovering under-recognized alterations in early-onset and H/L patient groups. The platform’s conversational interface, robust analytical engine, and harmonized integration of clinical-genomic data represent a scalable solution for precision oncology research and equitable biomarker discovery.

Future directions for AI-HOPE-MAPK include expansion to additional molecular pathways (e.g., TP53, TGF-β, WNT), integration of multi-omics (34) and spatial transcriptomics/proteomics (35), and deployment in prospective clinical trial design. Critically, our work supports the use of AI-powered tools to accelerate the identification of population-specific vulnerabilities, enhance translational research in diverse communities, and ultimately inform therapeutic decision-making for CRC patients who have historically been excluded from precision medicine pipelines. As the field moves toward increasingly data-driven, inclusive models of cancer care, platforms like AI-HOPE-MAPK will be essential in translating complexity into clinical action.

## Conclusion

AI-HOPE-MAPK represents a significant advancement in the application of artificial intelligence to precision oncology, enabling dynamic, population-aware exploration of MAPK pathway alterations in CRC. Through natural language-driven workflows, the platform integrates genomic and clinical data to uncover age– and ancestry-specific patterns that are often overlooked in traditional analyses. Our study demonstrates its utility in revealing differential mutation burdens, prognostic implications of low TMB MAPK-altered subtypes, and treatment-associated vulnerabilities in H/L EOCRC patients. These insights not only recapitulate known biology but also highlight novel disparities with clinical relevance. As the field advances toward inclusive and adaptive cancer care, AI-HOPE-MAPK offers a scalable solution for translating complex data into actionable knowledge—supporting efforts to personalize treatment, close equity gaps, and refine our molecular understanding of CRC across multiple patient populations.

## Data Availability

All data used in the present study is publicly available at https://www.cbioportal.org/ and https://genie.cbioportal.org. Additional data can be provided upon reasonable request to the authors.

---

**Figure S1.**
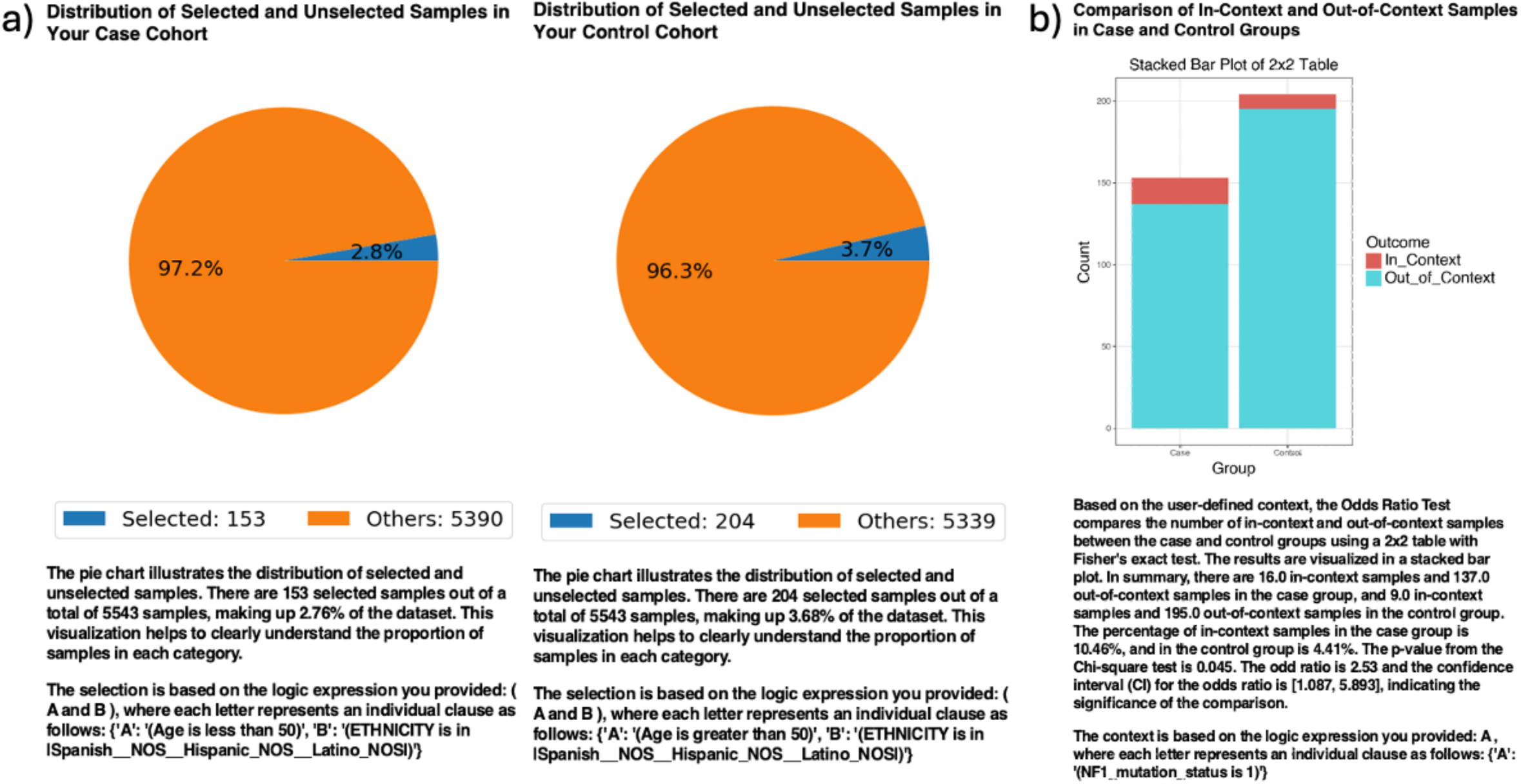
AI-HOPE-MAPK analysis of NF1 mutation frequency in early-vs. late-onset colorectal cancer among Hispanic/Latino patients. a) The pie charts display the proportion of selected samples in the case and control cohorts after natural language–based filtering using AI-HOPE-MAPK. The case cohort consists of 153 Hispanic/Latino patients diagnosed with early-onset colorectal cancer (EOCRC, under age 50), representing 2.8% of the harmonized dataset. The control cohort includes 204 Hispanic/Latino patients with late-onset colorectal cancer (LOCRC, over age 50), comprising 3.7% of the dataset. These charts visualize the relative representation of both groups used in downstream mutation analyses. b) A 2×2 contingency analysis evaluates the frequency of NF1 mutations between EOCRC and LOCRC Hispanic/Latino cohorts. The stacked bar plot shows the distribution of samples classified as “In_Context” (NF1 mutated) versus “Out_of_Context” (NF1 wild-type). NF1 mutations were present in 10.46% of EOCRC HL cases compared to 4.41% of LOCRC HL controls. The calculated odds ratio was 2.53 with a 95% confidence interval of [1.087, 5.893] and a p-value of 0.045, indicating a statistically significant enrichment of NF1 alterations in the early-onset group. These findings suggest that NF1 may play a role in the molecular etiology of EOCRC in Hispanic/Latino populations and highlight its potential relevance as a biomarker for age-related differences in CRC pathogenesis.

**Figure S2.**
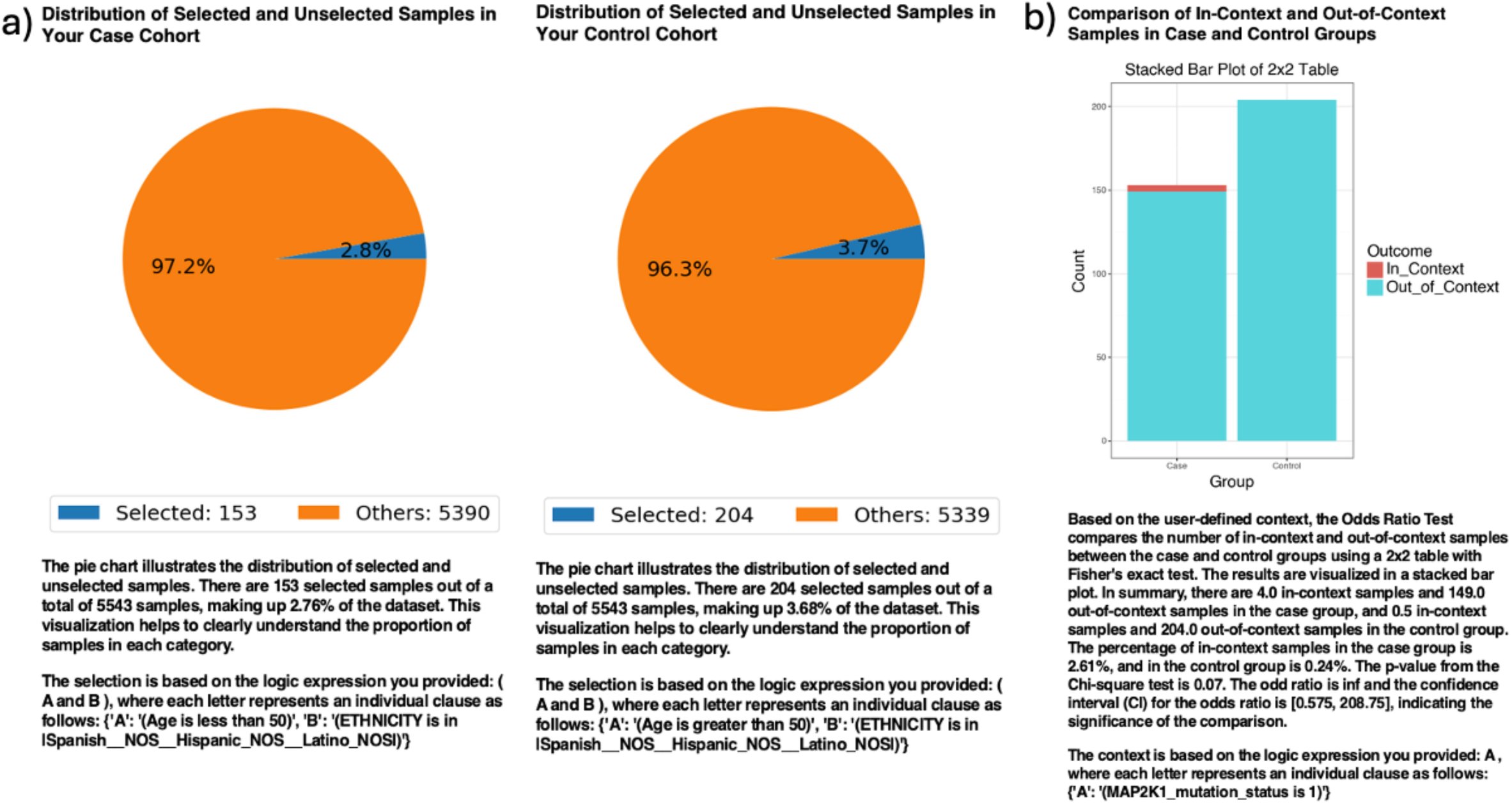
AI-HOPE-MAPK analysis of MAP2K1 mutation enrichment in early-versus late-onset colorectal cancer among Hispanic/Latino patients. a) Pie charts display the distribution of selected samples from the harmonized dataset following natural language–based filtering via AI-HOPE-MAPK. The case cohort includes 153 Hispanic/Latino patients diagnosed with early-onset colorectal cancer (EOCRC, under age 50), representing 2.8% of the dataset. The control cohort consists of 204 Hispanic/Latino patients with late-onset colorectal cancer (LOCRC, over age 50), comprising 3.7% of the dataset. These visualizations contextualize the relative sample proportions used for comparative genomic analysis. b) A 2×2 contingency analysis evaluates the presence of MAP2K1 mutations in EOCRC HL versus LOCRC HL groups. The stacked bar plot illustrates the number of “In_Context” (MAP2K1-mutated) and “Out_of_Context” (non-mutated) samples per group. MAP2K1 mutations were identified in 2.61% of EOCRC HL cases compared to just 0.24% of LOCRC HL controls. The odds ratio was estimated as infinite due to zero observed events in the control group, with a p-value of 0.07 and a 95% confidence interval ranging from 0.575 to 208.75. While the association did not reach conventional statistical significance, the magnitude of effect and low frequency of mutations in the older cohort highlight MAP2K1 as a potential driver gene in early-onset CRC among Hispanic/Latino patients. These results suggest the need for further validation in larger, ancestry-informed datasets.

**Figure S3.**
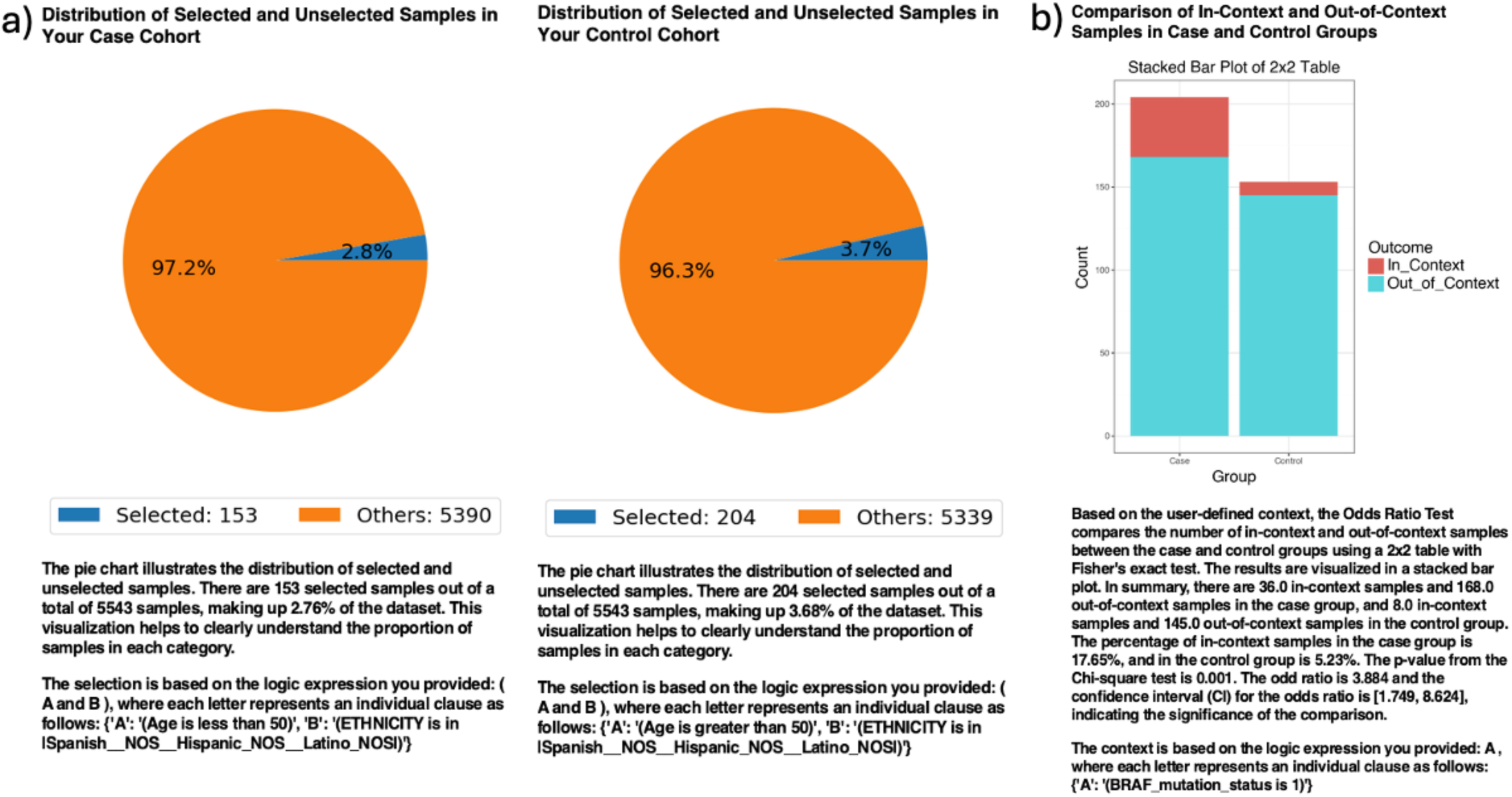
AI-HOPE-MAPK analysis of BRAF mutation prevalence in early-versus late-onset colorectal cancer among Hispanic/Latino patients. a) The pie charts depict the proportion of selected samples following natural language–based cohort filtering using AI-HOPE-MAPK. The early-onset colorectal cancer (EOCRC) Hispanic/Latino (HL) cohort includes 153 patients under age 50, representing 2.8% of the dataset. The late-onset colorectal cancer (LOCRC) HL cohort consists of 204 patients over age 50, comprising 3.7% of the dataset. These visualizations reflect the sample distributions used in the mutation frequency comparison. b) A 2×2 contingency analysis evaluates the frequency of BRAF mutations between LOCRC HL (case) and EOCRC HL (control) groups. The stacked bar plot shows “In_Context” (BRAF-mutated) and “Out_of_Context” (BRAF wild-type) samples within each group. BRAF mutations were identified in 17.65% of LOCRC HL cases compared to 5.23% of EOCRC HL cases. The resulting odds ratio was 3.88, with a statistically significant p-value of 0.001 and a 95% confidence interval of [1.749, 8.624]. These findings indicate that BRAF mutations are significantly enriched in the late-onset cohort, suggesting a possible age-associated molecular distinction in CRC pathogenesis within the Hispanic/Latino population.

**Figure S4.**
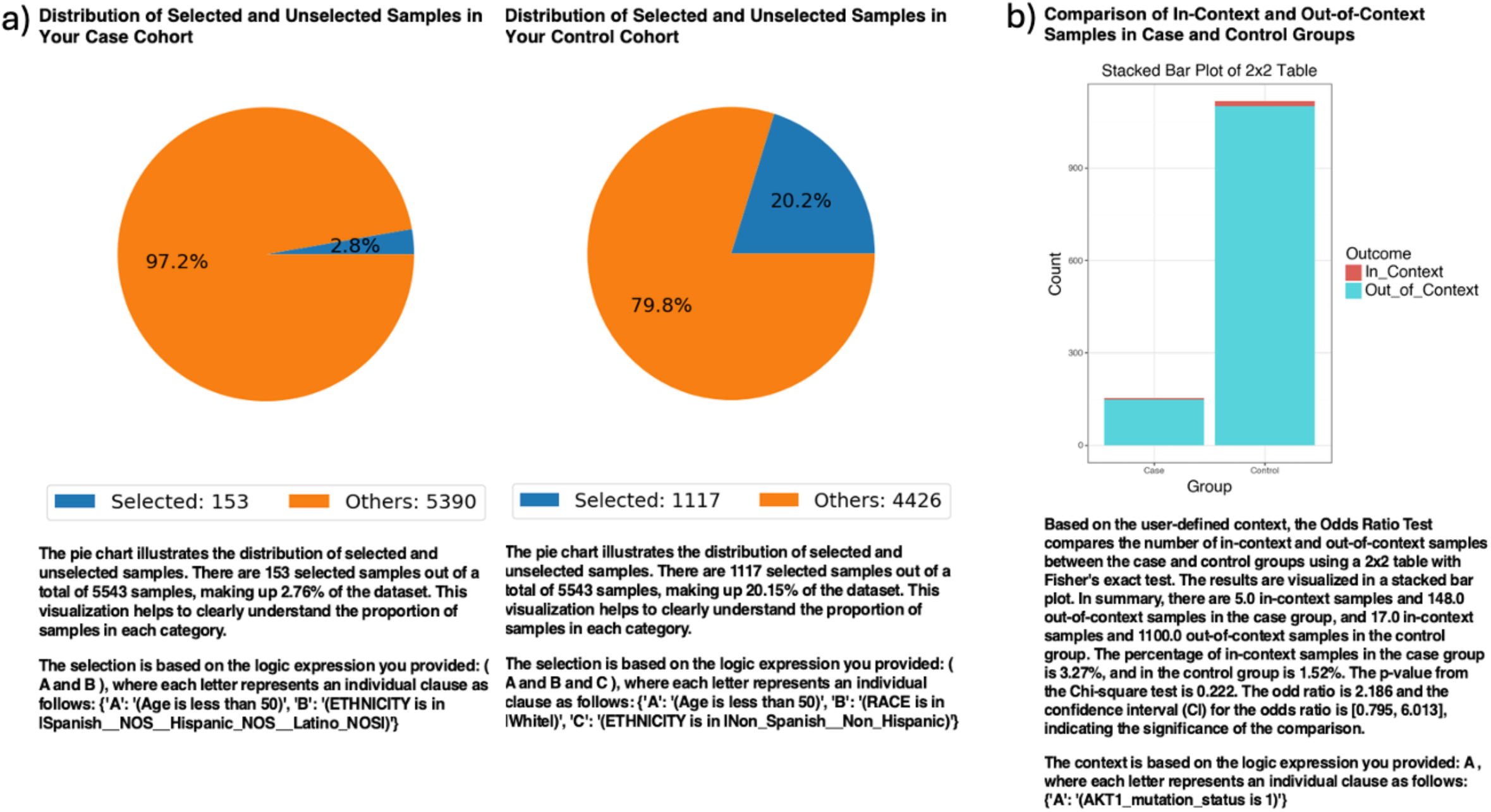
AI-HOPE-MAPK analysis of AKT1 mutation frequency in early-onset colorectal cancer among Hispanic/Latino and Non-Hispanic White patients. a) Pie charts illustrate the distribution of selected samples following natural language–driven filtering performed by AI-HOPE-MAPK. The case cohort comprises 153 Hispanic/Latino patients under age 50 diagnosed with early-onset colorectal cancer (EOCRC HL), representing 2.8% of the dataset. The control cohort includes 1,117 Non-Hispanic White (NHW) EOCRC patients under age 50, comprising 20.2% of the dataset. These proportions contextualize the groups used in subsequent comparative analyses. b) A 2×2 odds ratio analysis evaluates the frequency of AKT1 mutations between the EOCRC HL and EOCRC NHW groups. The stacked bar chart displays the number of “In_Context” (AKT1-mutated) and “Out_of_Context” (non-mutated) samples in each cohort. AKT1 mutations were observed in 3.27% of EOCRC HL patients and 1.52% of EOCRC NHW patients. The calculated odds ratio was 2.19, with a 95% confidence interval of [0.795, 6.013] and a p-value of 0.222. Although the results did not reach statistical significance, the observed trend suggests a potential enrichment of AKT1 mutations in the Hispanic/Latino EOCRC population. Further investigation in larger, ancestry-stratified cohorts is warranted to validate the clinical relevance of this finding.

**Figure S5.**
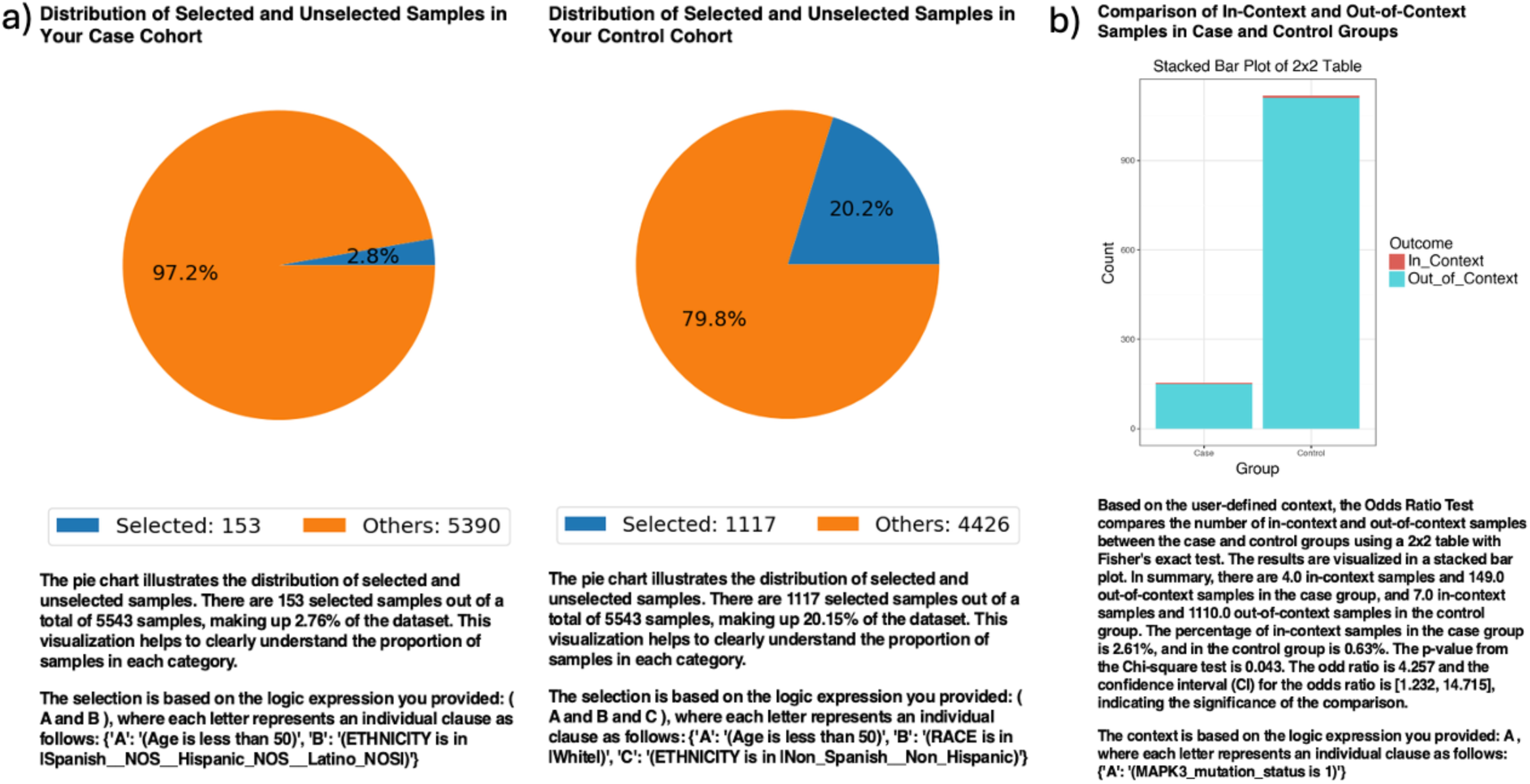
AI-HOPE-MAPK analysis of MAPK3 mutation frequency in early-onset colorectal cancer among Hispanic/Latino and Non-Hispanic White patients. a) Pie charts show the distribution of selected samples after AI-HOPE-MAPK’s natural language–guided filtering of the harmonized dataset. The case cohort includes 153 early-onset colorectal cancer (EOCRC) patients under age 50 identified as Hispanic/Latino (HL), representing 2.8% of the dataset. The control cohort consists of 1,117 EOCRC patients identified as Non-Hispanic White (NHW), representing 20.2% of the total sample set. These visuals reflect the cohort definitions used in the comparative genomic analysis. b) A 2×2 contingency analysis compares the prevalence of MAPK3 mutations between EOCRC HL and EOCRC NHW groups. The stacked bar plot presents the number of samples with MAPK3 mutations (“In_Context”) and those without (“Out_of_Context”) in each cohort. MAPK3 mutations were detected in 2.61% of EOCRC HL cases and 0.63% of EOCRC NHW cases. The calculated odds ratio was 4.26, with a p-value of 0.043 and a 95% confidence interval of [1.232, 14.715], indicating statistical significance. These results suggest that MAPK3 mutations are significantly enriched in the Hispanic/Latino early-onset CRC population. The findings support further investigation into MAPK3 as a potential biomarker of ancestry-associated molecular variation and therapeutic relevance in EOCRC.

**Figure S6.**
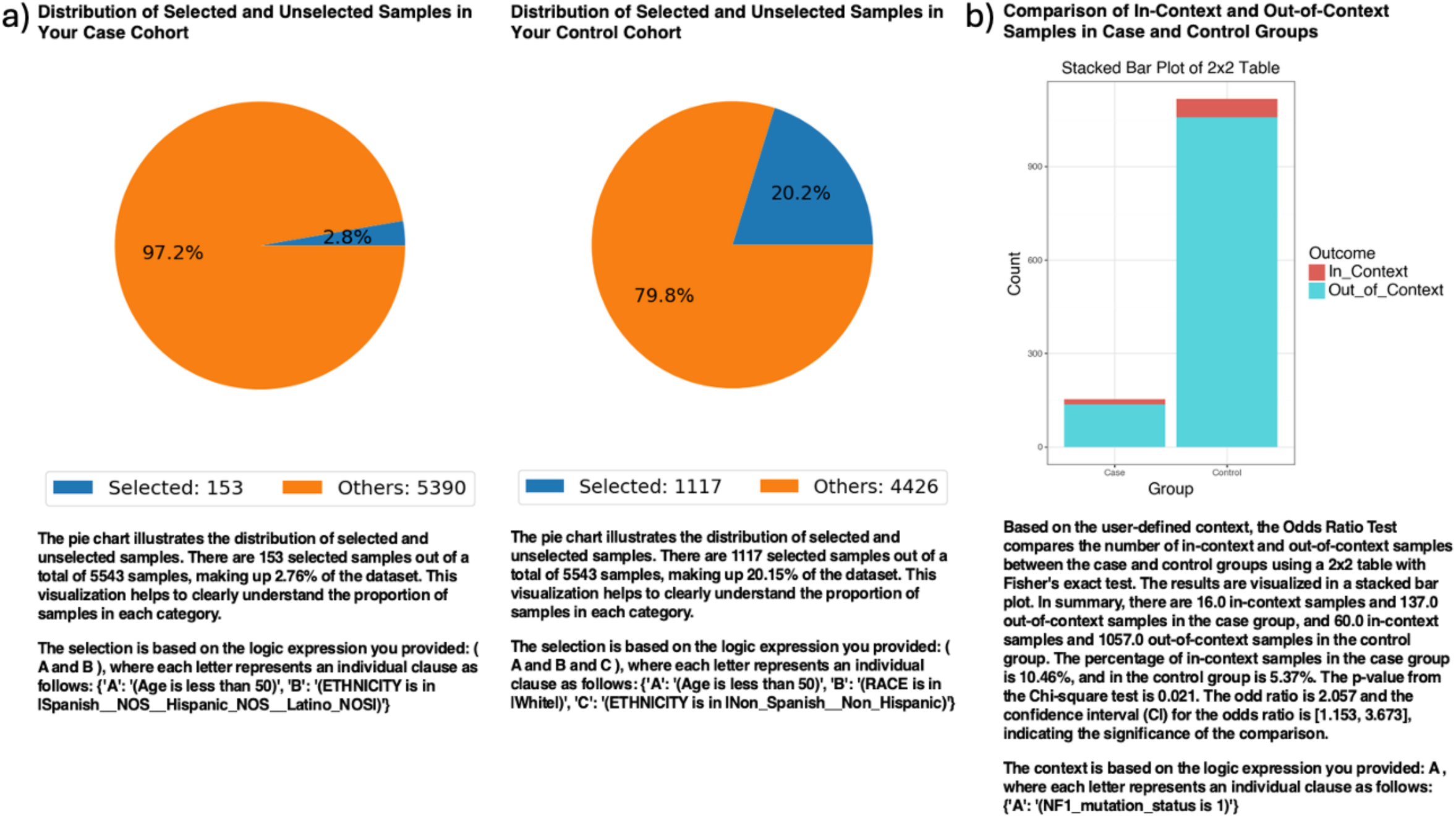
AI-HOPE-MAPK analysis of NF1 mutation prevalence in early-onset colorectal cancer among Hispanic/Latino and Non-Hispanic White populations. a) Pie charts visualize the proportion of selected samples in each cohort after automated filtering using AI-HOPE-MAPK’s natural language–based cohort construction. The case cohort includes 153 Hispanic/Latino patients under age 50 with early-onset colorectal cancer (EOCRC HL), representing 2.8% of the dataset. The control cohort comprises 1,117 Non-Hispanic White (NHW) EOCRC patients, accounting for 20.2% of the dataset. These visual summaries provide a clear view of the sample composition used in downstream mutation comparisons. b) A 2×2 odds ratio analysis examines the frequency of NF1 mutations between the EOCRC HL and EOCRC NHW cohorts. The bar plot illustrates the number of samples with NF1 mutations (“In_Context”) versus those without (“Out_of_Context”). NF1 mutations were identified in 10.46% of EOCRC HL samples compared to 5.37% of EOCRC NHW samples. The odds ratio was 2.06, with a 95% confidence interval of [1.153, 3.673] and a statistically significant p-value of 0.021. These findings suggest a significant enrichment of NF1 mutations in the Hispanic/Latino EOCRC cohort, underscoring the potential role of NF1 as a molecular driver of ancestry-associated disparities in early-onset CRC.

**Figure S7.**
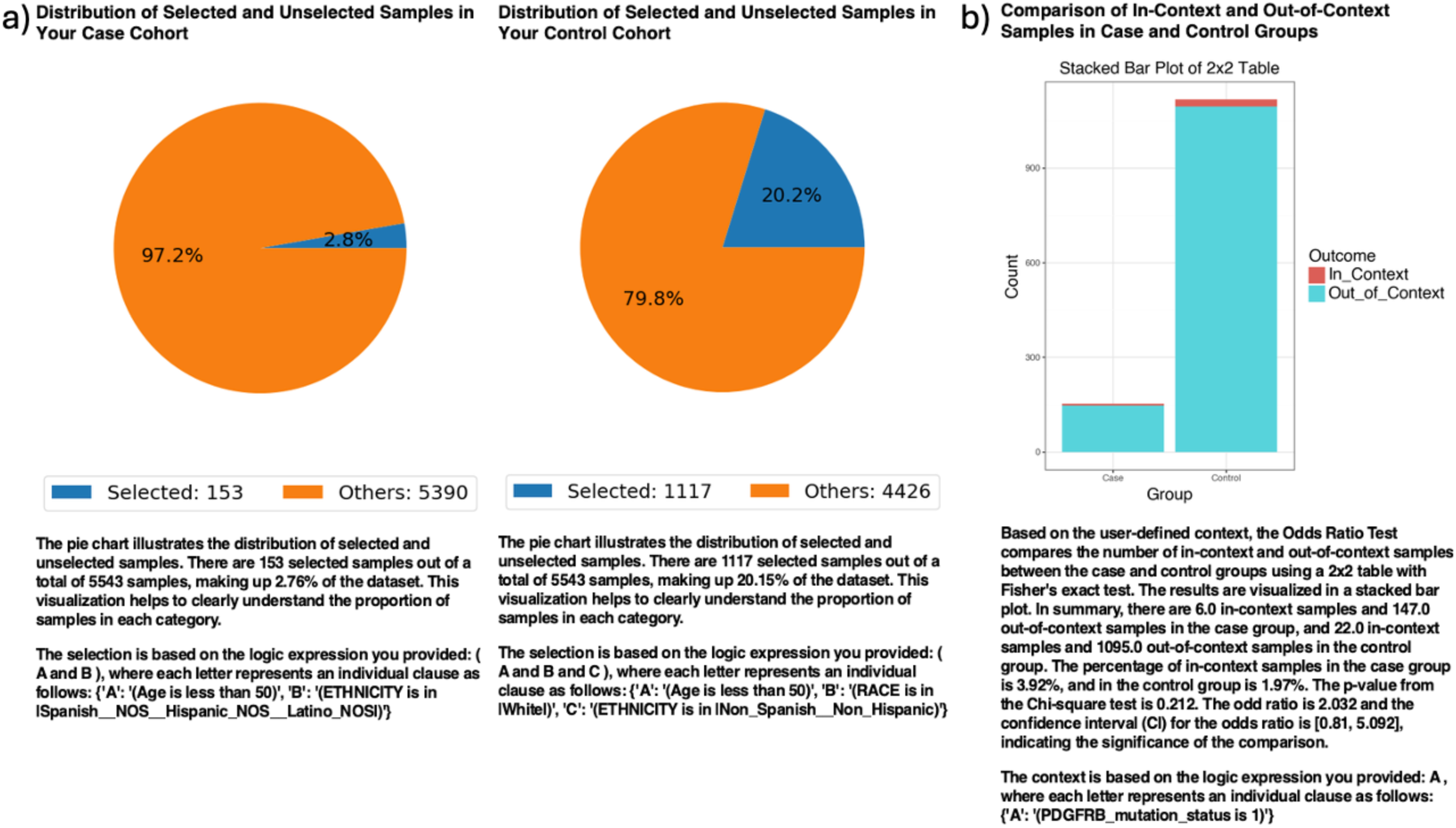
AI-HOPE-MAPK analysis of PDGFRB mutation prevalence in early-onset colorectal cancer among Hispanic/Latino (H/L) and Non-Hispanic White (NHW) patients. a) Pie charts illustrate the proportion of selected samples in each cohort following automated, natural language–guided filtering by AI-HOPE-MAPK. The case cohort consists of 153 early-onset colorectal cancer (EOCRC) patients under age 50 who self-identified as H/L, representing 2.8% of the dataset. The control cohort includes 1,117 EOCRC patients under age 50 who identified as NHW, comprising 20.2% of the total samples. These visualizations provide context for the cohort distribution used in mutation frequency comparisons. b) A 2×2 odds ratio analysis evaluates the frequency of PDGFRB mutations across the EOCRC HL and EOCRC NHW groups. The stacked bar plot shows the number of samples classified as “In_Context” (PDGFRB-mutated) versus “Out_of_Context” (non-mutated). PDGFRB mutations were found in 3.92% of HL samples and 1.97% of NHW samples. The calculated odds ratio was 2.03, with a p-value of 0.212 and a 95% confidence interval of [0.81, 5.092], indicating that the observed difference was not statistically significant. Nonetheless, this trend suggests a possible enrichment of PDGFRB mutations in the Hispanic/Latino early-onset population. These results warrant further investigation in larger, ancestry-informed cohorts to assess the potential role of PDGFRB in population-specific CRC biology.

**Figure S8.**
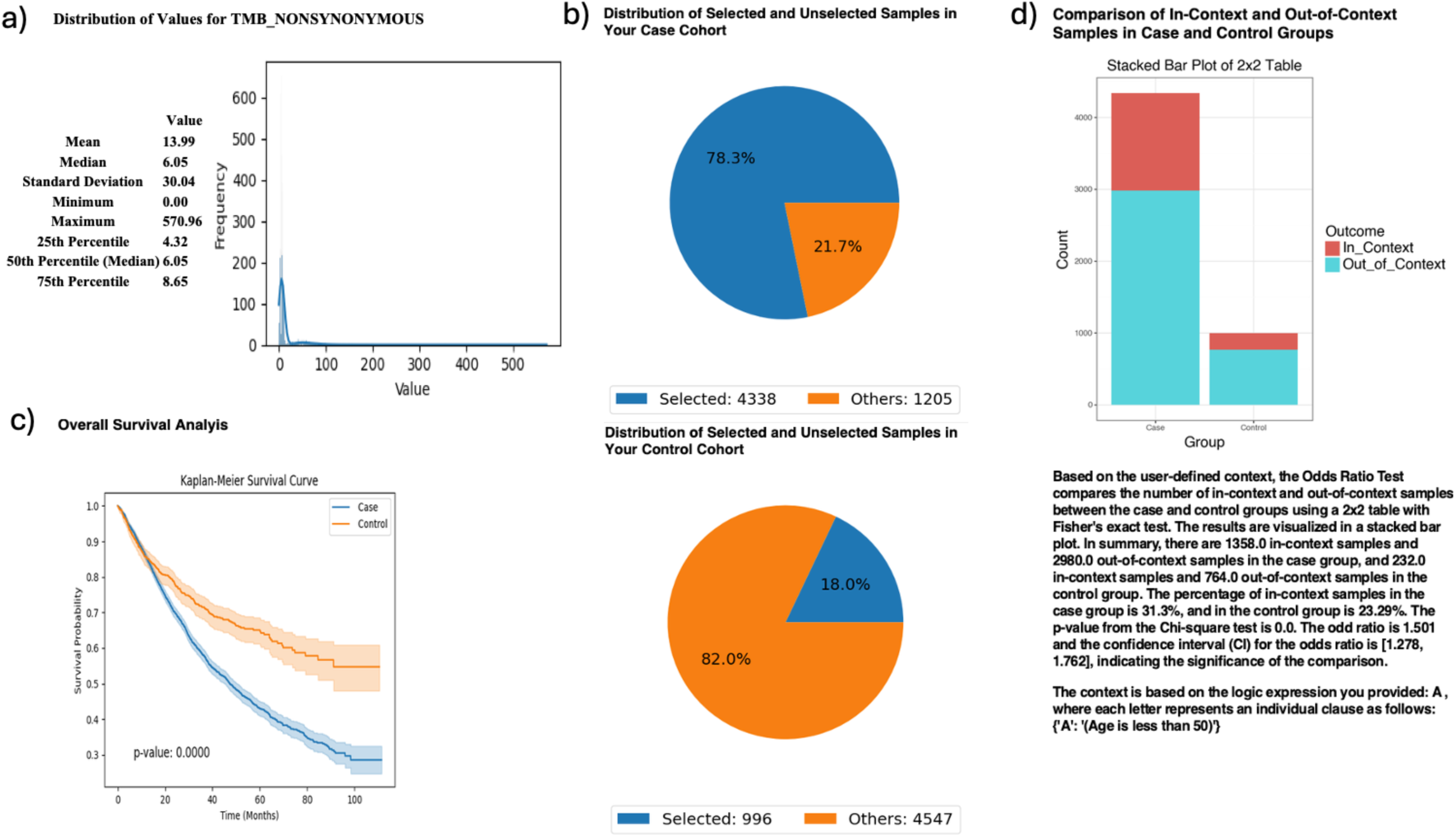
AI-HOPE-MAPK evaluation of survival outcomes in MAPK-altered colorectal cancer stratified by tumor mutational burden (TMB) and age. This figure demonstrates the use of AI-HOPE-MAPK to assess survival differences and mutational context among colorectal cancer (CRC) patients stratified by MAPK pathway alteration status and tumor mutational burden (TMB). The case cohort consists of patients with MAPK pathway alterations and low TMB (<10 mutations/Mb), while the control cohort includes those with no MAPK alterations and high TMB (>10 mutations/Mb). The analysis further considers age (<50 years) as an odds ratio test context. a) A histogram illustrates the distribution of TMB_NONSYNONYMOUS values across the dataset, showing a right-skewed distribution with a median of 6.05 and a long tail extending beyond 500. This visualization supports the selection of 10 mutations/Mb as a threshold for defining high vs. low TMB. b) Pie charts display the selection proportions for each cohort. The case group includes 4,338 MAPK-altered, low-TMB samples (78.3% of selected), while the control cohort includes 996 high-TMB, MAPK-wildtype samples (18.0% of selected), highlighting a substantial representation of MAPK-altered cases. c) Kaplan-Meier survival curves compare overall survival between the two groups. Patients in the MAPK-altered, low-TMB case group experienced significantly poorer survival compared to controls, with a p-value < 0.0001. Shaded confidence intervals emphasize the divergence in survival trajectories. d) A 2×2 odds ratio analysis was conducted to evaluate whether MAPK-altered, low-TMB tumors are enriched among younger CRC patients. Among the case cohort, 31.8% of samples belonged to patients under 50, compared to 23.29% in the control group. The resulting odds ratio was 1.501 (95% CI: 1.278–1.762; p < 0.001), indicating a statistically significant enrichment of MAPK pathway alterations in younger CRC patients with low TMB. These results suggest that MAPK-driven, low-TMB tumors may represent a distinct molecular subtype with worse prognosis in early-onset colorectal cancer.

**Figure S9.**
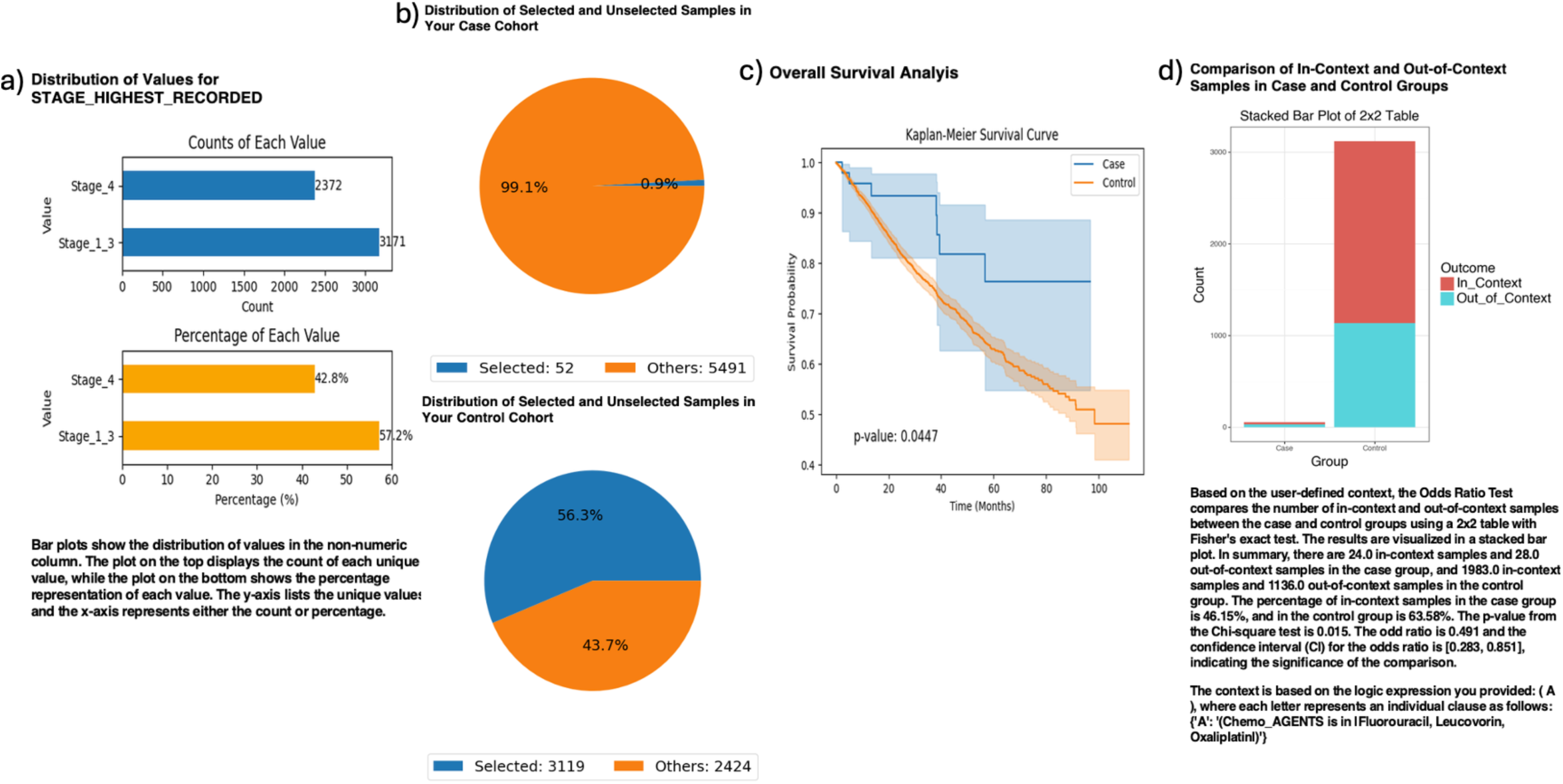
AI-HOPE-MAPK analysis of stage I–III colorectal cancer patients treated with FOLFOX, stratified by MAP2K1 mutation status. This figure illustrates the application of AI-HOPE-MAPK to evaluate clinical outcomes and treatment context among colorectal cancer (CRC) patients with early-stage disease (Stage I–III), comparing those with MAP2K1 mutations (case cohort) to MAP2K1 wild-type patients (control cohort), all of whom received standard FOLFOX chemotherapy (Fluorouracil, Leucovorin, Oxaliplatin). a) Bar plots summarize the distribution of the highest stage recorded across the dataset, revealing that 57.2% of cases were Stage I–III, while 42.8% were Stage IV. This justified the selection of patients with Stage I–III disease for focused comparison. b) Pie charts show cohort selection proportions. The case cohort includes 52 MAP2K1-mutant Stage I–III samples (0.9% of the dataset), while the control cohort includes 3,119 wild-type counterparts (56.3%). These visualizations underscore the rarity of MAP2K1 mutations in early-stage CRC. c) Kaplan-Meier analysis compares overall survival between the two groups, revealing a statistically significant difference (p = 0.0447). Patients with MAP2K1 mutations had worse survival outcomes, as indicated by the divergence of the curves and non-overlapping confidence intervals. d) A 2×2 odds ratio analysis was performed to assess whether MAP2K1-mutant patients were less likely to receive FOLFOX. Among the case cohort, 46.15% were treated with FOLFOX compared to 63.55% in the control group, yielding an odds ratio of 0.491 (95% CI: [0.283, 0.851]; p = 0.015). These results suggest a significant underrepresentation of FOLFOX exposure among MAP2K1-mutant early-stage patients, raising the possibility of treatment divergence or biological resistance mechanisms requiring further exploration.

## References

1. Sung H, Ferlay J, Siegel RL, Laversanne M, Soerjomataram I, Jemal A, Bray F. Global Cancer Statistics 2020: GLOBOCAN Estimates of Incidence and Mortality Worldwide for 36 Cancers in 185 Countries. CA Cancer J Clin. 2021 May;71(3):209–249. doi: 10.3322/caac.21660. Epub 2021 Feb 4. PMID: 33538338.

2. Siegel RL, Miller KD, Wagle NS, Jemal A. Cancer statistics, 2023. CA Cancer J Clin. 2023 Jan;73(1):17–48. doi: 10.3322/caac.21763. PMID: 36633525.

3. Siegel RL, Fedewa SA, Anderson WF, Miller KD, Ma J, Rosenberg PS, Jemal A. Colorectal Cancer Incidence Patterns in the United States, 1974-2013. J Natl Cancer Inst. 2017 Aug 1;109(8):djw322. doi: 10.1093/jnci/djw322. PMID: 28376186; PMCID: PMC6059239.

4. Araghi M, Soerjomataram I, Jenkins M, Brierley J, Morris E, Bray F, Arnold M. Global trends in colorectal cancer mortality: projections to the year 2035. Int J Cancer. 2019 Jun 15;144(12):2992–3000. doi: 10.1002/ijc.32055. Epub 2019 Jan 8. PMID: 30536395.

5. Monge C, Waldrup B, Carranza FG, Velazquez-Villarreal E. Ethnicity-Specific Molecular Alterations in MAPK and JAK/STAT Pathways in Early-Onset Colorectal Cancer. Cancers (Basel). 2025 Mar 25;17(7):1093. doi: 10.3390/cancers17071093. PMID: 40227607; PMCID: PMC11988162.

6. Patel SG, Karlitz JJ, Yen T, Lieu CH, Boland CR. Comparison of trends in early-onset colorectal cancer in North America and Europe – Authors’ reply. Lancet Gastroenterol Hepatol. 2022 Jun;7(6):506. doi: 10.1016/S2468-1253(22)00123-6. PMID: 35550052.

7. Koblinski J, Jandova J, Nfonsam V. Disparities in incidence of early– and late-onset colorectal cancer between Hispanics and Whites: A 10-year SEER database study. Am J Surg. 2018 Apr;215(4):581–585. doi: 10.1016/j.amjsurg.2017.03.035. Epub 2017 Mar 31. PMID: 28388972.

8. De Carvalho TC, Borges AKDM, Da Silva IF. Incidence of Colorectal Cancer in Selected Countries of Latin America: Age-Period-Cohort Effect. Asian Pac J Cancer Prev. 2020 Nov 1;21(11):3421–3428. doi: 10.31557/APJCP.2020.21.11.3421. PMID: 33247704; PMCID: PMC8033126.

9. Monge C, Waldrup B, Carranza FG, Velazquez-Villarreal E. WNT and TGF-Beta Pathway Alterations in Early-Onset Colorectal Cancer Among Hispanic/Latino Populations. Cancers (Basel). 2024 Nov 21;16(23):3903. doi: 10.3390/cancers16233903. PMID: 39682092; PMCID: PMC11639970.

10. Muller C, Ihionkhan E, Stoffel EM, Kupfer SS. Disparities in Early-Onset Colorectal Cancer. Cells. 2021 Apr 26;10(5):1018. doi: 10.3390/cells10051018. PMID: 33925893; PMCID: PMC8146231.

11. Demb J, Gomez SL, Canchola AJ, Qian A, Murphy JD, Winn RA, Banegas MP, Gupta S, Martinez ME. Racial and Ethnic Variation in Survival in Early-Onset Colorectal Cancer. JAMA Netw Open. 2024 Nov 4;7(11):e2446820. doi: 10.1001/jamanetworkopen.2024.46820. PMID: 39576642; PMCID: PMC11584933.

12. Wolf AMD, Fontham ETH, Church TR, Flowers CR, Guerra CE, LaMonte SJ, Etzioni R, McKenna MT, Oeffinger KC, Shih YT, Walter LC, Andrews KS, Brawley OW, Brooks D, Fedewa SA, Manassaram-Baptiste D, Siegel RL, Wender RC, Smith RA. Colorectal cancer screening for average-risk adults: 2018 guideline update from the American Cancer Society. CA Cancer J Clin. 2018 Jul;68(4):250–281. doi: 10.3322/caac.21457. Epub 2018 May 30. PMID: 29846947.

13. Holowatyj AN, Gigic B, Herpel E, Scalbert A, Schneider M, Ulrich CM; MetaboCCC Consortium; ColoCare Study. Distinct Molecular Phenotype of Sporadic Colorectal Cancers Among Young Patients Based on Multiomics Analysis. Gastroenterology. 2020 Mar;158(4):1155–1158.e2. doi: 10.1053/j.gastro.2019.11.012. Epub 2019 Nov 13. PMID: 31730769; PMCID: PMC7291587.

14. Yaeger R, Chatila WK, Lipsyc MD, Hechtman JF, Cercek A, Sanchez-Vega F, Jayakumaran G, Middha S, Zehir A, Donoghue MTA, You D, Viale A, Kemeny N, Segal NH, Stadler ZK, Varghese AM, Kundra R, Gao J, Syed A, Hyman DM, Vakiani E, Rosen N, Taylor BS, Ladanyi M, Berger MF, Solit DB, Shia J, Saltz L, Schultz N. Clinical Sequencing Defines the Genomic Landscape of Metastatic Colorectal Cancer. Cancer Cell. 2018 Jan 8;33(1):125–136.e3. doi: 10.1016/j.ccell.2017.12.004. PMID: 29316426; PMCID: PMC5765991.

15. Llosa NJ, Cruise M, Tam A, Wicks EC, Hechenbleikner EM, Taube JM, Blosser RL, Fan H, Wang H, Luber BS, Zhang M, Papadopoulos N, Kinzler KW, Vogelstein B, Sears CL, Anders RA, Pardoll DM, Housseau F. The vigorous immune microenvironment of microsatellite instable colon cancer is balanced by multiple counter-inhibitory checkpoints. Cancer Discov. 2015 Jan;5(1):43–51. doi: 10.1158/2159-8290.CD-14-0863. Epub 2014 Oct 30. PMID: 25358689; PMCID: PMC4293246.

16. Thorsson V, Gibbs DL, Brown SD, Wolf D, Bortone DS, Ou Yang TH, Porta-Pardo E, Gao GF, Plaisier CL, Eddy JA, Ziv E, Culhane AC, Paull EO, Sivakumar IKA, Gentles AJ, Malhotra R, Farshidfar F, Colaprico A, Parker JS, Mose LE, Vo NS, Liu J, Liu Y, Rader J, Dhankani V, Reynolds SM, Bowlby R, Califano A, Cherniack AD, Anastassiou D, Bedognetti D, Mokrab Y, Newman AM, Rao A, Chen K, Krasnitz A, Hu H, Malta TM, Noushmehr H, Pedamallu CS, Bullman S, Ojesina AI, Lamb A, Zhou W, Shen H, Choueiri TK, Weinstein JN, Guinney J, Saltz J, Holt RA, Rabkin CS; Cancer Genome Atlas Research Network; Lazar AJ, Serody JS, Demicco EG, Disis ML, Vincent BG, Shmulevich I. The Immune Landscape of Cancer. Immunity. 2018 Apr 17;48(4):812–830.e14. doi: 10.1016/j.immuni.2018.03.023. Epub 2018 Apr 5. Erratum in: Immunity. 2019 Aug 20;51(2):411-412. doi: 10.1016/j.immuni.2019.08.004. PMID: 29628290; PMCID: PMC5982584.

17. Baba Y, Huttenhower C, Nosho K, Tanaka N, Shima K, Hazra A, Schernhammer ES, Hunter DJ, Giovannucci EL, Fuchs CS, Ogino S. Epigenomic diversity of colorectal cancer indicated by LINE-1 methylation in a database of 869 tumors. Mol Cancer. 2010 May 27;9:125. doi: 10.1186/1476-4598-9-125. PMID: 20507599; PMCID: PMC2892454.

18. Dhillon AS, Hagan S, Rath O, Kolch W. MAP kinase signalling pathways in cancer. Oncogene. 2007 May 14;26(22):3279–90. doi: 10.1038/sj.onc.1210421. PMID: 17496922.

19. Hymowitz SG, Malek S. Targeting the MAPK Pathway in RAS Mutant Cancers. Cold Spring Harb Perspect Med. 2018 Nov 1;8(11):a031492. doi: 10.1101/cshperspect.a031492. PMID: 29440321; PMCID: PMC6211377.

20. Burotto M, Chiou VL, Lee JM, Kohn EC. The MAPK pathway across different malignancies: a new perspective. Cancer. 2014 Nov 15;120(22):3446–56. doi: 10.1002/cncr.28864. Epub 2014 Jun 19. PMID: 24948110; PMCID: PMC4221543.

21. Koveitypour Z, Panahi F, Vakilian M, Peymani M, Seyed Forootan F, Nasr Esfahani MH, Ghaedi K. Signaling pathways involved in colorectal cancer progression. Cell Biosci. 2019 Dec 2;9:97. doi: 10.1186/s13578-019-0361-4. PMID: 31827763; PMCID: PMC6889432.

22. Fang JY, Richardson BC. The MAPK signalling pathways and colorectal cancer. Lancet Oncol. 2005 May;6(5):322–7. doi: 10.1016/S1470-2045(05)70168-6. PMID: 15863380.

23. Corcoran RB, Ebi H, Turke AB, Coffee EM, Nishino M, Cogdill AP, Brown RD, Della Pelle P, Dias-Santagata D, Hung KE, Flaherty KT, Piris A, Wargo JA, Settleman J, Mino-Kenudson M, Engelman JA. EGFR-mediated re-activation of MAPK signaling contributes to insensitivity of BRAF mutant colorectal cancers to RAF inhibition with vemurafenib. Cancer Discov. 2012 Mar;2(3):227–35. doi: 10.1158/2159-8290.CD-11-0341. Epub 2012 Jan 16. PMID: 22448344; PMCID: PMC3308191.

24. Bahar ME, Kim HJ, Kim DR. Targeting the RAS/RAF/MAPK pathway for cancer therapy: from mechanism to clinical studies. Signal Transduct Target Ther. 2023 Dec 18;8(1):455. doi: 10.1038/s41392-023-01705-z. PMID: 38105263; PMCID: PMC10725898.

25. Kikuchi H, Pino MS, Zeng M, Shirasawa S, Chung DC. Oncogenic KRAS and BRAF differentially regulate hypoxia-inducible factor-1alpha and –2alpha in colon cancer. Cancer Res. 2009 Nov 1;69(21):8499–506. doi: 10.1158/0008-5472.CAN-09-2213. Epub 2009 Oct 20. PMID: 19843849; PMCID: PMC2811371.

26. De Roock W, Claes B, Bernasconi D, De Schutter J, Biesmans B, Fountzilas G, Kalogeras KT, Kotoula V, Papamichael D, Laurent-Puig P, Penault-Llorca F, Rougier P, Vincenzi B, Santini D, Tonini G, Cappuzzo F, Frattini M, Molinari F, Saletti P, De Dosso S, Martini M, Bardelli A, Siena S, Sartore-Bianchi A, Tabernero J, Macarulla T, Di Fiore F, Gangloff AO, Ciardiello F, Pfeiffer P, Qvortrup C, Hansen TP, Van Cutsem E, Piessevaux H, Lambrechts D, Delorenzi M, Tejpar S. Effects of KRAS, BRAF, NRAS, and PIK3CA mutations on the efficacy of cetuximab plus chemotherapy in chemotherapy-refractory metastatic colorectal cancer: a retrospective consortium analysis. Lancet Oncol. 2010 Aug;11(8):753–62. doi: 10.1016/S1470-2045(10)70130-3. Epub 2010 Jul 8. PMID: 20619739.

27. Corcoran RB, Ebi H, Turke AB, Coffee EM, Nishino M, Cogdill AP, Brown RD, Della Pelle P, Dias-Santagata D, Hung KE, Flaherty KT, Piris A, Wargo JA, Settleman J, Mino-Kenudson M, Engelman JA. EGFR-mediated re-activation of MAPK signaling contributes to insensitivity of BRAF mutant colorectal cancers to RAF inhibition with vemurafenib. Cancer Discov. 2012 Mar;2(3):227–35. doi: 10.1158/2159-8290.CD-11-0341. Epub 2012 Jan 16. PMID: 22448344; PMCID: PMC3308191.

28. Cancer Genome Atlas Network. Comprehensive molecular characterization of human colon and rectal cancer. Nature. 2012 Jul 18;487(7407):330–7. doi: 10.1038/nature11252. PMID: 22810696; PMCID: PMC3401966.

29. Guinney J, Dienstmann R, Wang X, de Reyniès A, Schlicker A, Soneson C, Marisa L, Roepman P, Nyamundanda G, Angelino P, Bot BM, Morris JS, Simon IM, Gerster S, Fessler E, De Sousa E, Melo F, Missiaglia E, Ramay H, Barras D, Homicsko K, Maru D, Manyam GC, Broom B, Boige V, Perez-Villamil B, Laderas T, Salazar R, Gray JW, Hanahan D, Tabernero J, Bernards R, Friend SH, Laurent-Puig P, Medema JP, Sadanandam A, Wessels L, Delorenzi M, Kopetz S, Vermeulen L, Tejpar S. The consensus molecular subtypes of colorectal cancer. Nat Med. 2015 Nov;21(11):1350–6. doi: 10.1038/nm.3967. Epub 2015 Oct 12. PMID: 26457759; PMCID: PMC4636487.

30. Dienstmann R, Vermeulen L, Guinney J, Kopetz S, Tejpar S, Tabernero J. Consensus molecular subtypes and the evolution of precision medicine in colorectal cancer. Nat Rev Cancer. 2017 Feb;17(2):79–92. doi: 10.1038/nrc.2016.126. Epub 2017 Jan 4. Erratum in: Nat Rev Cancer. 2017 Mar 23;17(4):268. doi: 10.1038/nrc.2017.24. PMID: 28050011.

31. Ahronian LG, Sennott EM, Van Allen EM, Wagle N, Kwak EL, Faris JE, Godfrey JT, Nishimura K, Lynch KD, Mermel CH, Lockerman EL, Kalsy A, Gurski JM Jr, Bahl S, Anderka K, Green LM, Lennon NJ, Huynh TG, Mino-Kenudson M, Getz G, Dias-Santagata D, Iafrate AJ, Engelman JA, Garraway LA, Corcoran RB. Clinical Acquired Resistance to RAF Inhibitor Combinations in BRAF-Mutant Colorectal Cancer through MAPK Pathway Alterations. Cancer Discov. 2015 Apr;5(4):358–67. doi: 10.1158/2159-8290.CD-14-1518. Epub 2015 Feb 11. PMID: 25673644; PMCID: PMC4390490.

32. Saklatvala J. The p38 MAP kinase pathway as a therapeutic target in inflammatory disease. Curr Opin Pharmacol. 2004 Aug;4(4):372–7. doi: 10.1016/j.coph.2004.03.009. PMID: 15251131.

33. Cohen R, Cervera P, Svrcek M, Pellat A, Dreyer C, de Gramont A, André T. BRAF-Mutated Colorectal Cancer: What Is the Optimal Strategy for Treatment? Curr Treat Options Oncol. 2017 Feb;18(2):9. doi: 10.1007/s11864-017-0453-5. PMID: 28214977.

34. Waldrup B, Carranza F, Jin Y, Amzaleg Y, Postel M, Craig DW, Carpten JD, Salhia B, Ricker CN, Culver JO, Chavez CE, Stern MC, Baezconde-Garbanati L, Lenz HJ, Velazquez-Villarreal EI. Integrative multi-omics profiling of colorectal cancer from a Hispanic/Latino cohort of patients. medRxiv [Preprint]. 2024 Nov 15:2024.11.03.24316599. doi: 10.1101/2024.11.03.24316599. PMID: 39606335; PMCID: PMC11601710.

35. Carranza FG, Diaz FC, Ninova M, Velazquez-Villarreal E. Current state and future prospects of spatial biology in colo-rectal cancer. Front Oncol. 2024 Dec 3;14:1513821. doi: 10.3389/fonc.2024.1513821. PMID: 39711954; PMCID: PMC11660798.

